# Determinants of Adherence to Cervical Cancer Prevention Strategies Among Socially Diverse Middle-Aged Women

**DOI:** 10.64898/2026.02.04.26345545

**Authors:** Natalia Marin, Blanca Sarzo, Raul Beneyto, Oihane Alvarez, Esther Gracia, Marisa Estarlich, Sabrina Llop, Ana Molina-Barceló, Ignacio G. Bravo, Reem Abumallouh, Maria-Jose Lopez-Espinosa

**Affiliations:** Foundation for the Promotion of Health and Biomedical Research in the Valencian Region, FISABIO-Public Health, Valencia, Spain; Spanish Consortium for Research on Epidemiology and Public Health (CIBERESP), Madrid, Spain; Faculty of Nursing and Chiropody, Universitat de València (UV), Valencia, Spain; Epidemiology and Environmental Health Joint Research Unit, FISABIO-Universitat Jaume I–Universitat de València, Valencia, Spain; Department of Applied Statistics and Operational Research, and Quality, Universitat Politècnica de València (UPV), Valencia, Spain; Department of Social Psychology, Universitat de València (UV), Valencia, Spain; IDOCAL Research Institute, Valencia, Spain; Laboratory MIVEGEC (CNRS, IRD, University of Montpellier), French National Centre for Scientific Research, Montpellier, France

**Keywords:** cervical cancer prevention, middle-aged women, underprivileged women, female sex worker, determinants

## Abstract

Despite the availability of vaccinations and screenings, cervical cancer (CC) remains a major global health challenge. Understanding the factors influencing adherence to CC prevention strategies is essential, particularly among overlooked groups such as middle-aged and/or underprivileged women. For this purpose, a cross-sectional study was conducted in a population comprising 379 women (aged 37–64 years, recruitment: 2019–2023) from Valencia (Spain), representing diverse socioeconomic profiles, including female sex workers (FSWs; n = 46). Data were analysed using multivariate logistic regression models. Overall, 88.65% of participants adhered to CCS recommendations and 22.16% reported past-year condom use, compared with 86.96% and 76.09%, respectively, among FSWs. In the overall population, CCS adherence was significantly associated with higher education and marginally associated with a history of candidiasis, non-condom contraceptive use in the past year, and having more children and sexual partners. Higher condom use was associated with lower alcohol consumption, being single, non-Spanish origin, not being postmenopausal, use of vaginal health products, and not previous recurrent urinary tract infections. Marginally significant positive associations were also observed for the absence of chronic disease and a history of sexually transmitted infections. Among FSWs, earlier menarche age and lower gravidity were marginally associated with higher CCS adherence, while older age at entry into prostitution and earlier sexual debut were associated with condom use, the latter marginally significant. Overall, our findings highlight the importance of further research into underprivileged groups such as midlife women across socioeconomic strata.

**Novelty and Impact:** First study to assess 28 determinants of CC prevention in middle-aged women from diverse socioeconomic backgrounds, an often overlooked group. Key factors were education, alcohol consumption, relationship status, birth country, menopausal status, vaginal product use, and recurrent UTIs. Weaker associations were found for candidiasis history, non-condom contraception, number of children and sexual partners, STI history, and chronic disease. Our findings underscore the need for targeted public-health interventions to support the WHO strategy to eliminate CC.

## 1. INTRODUCTION

Cervical cancer (CC) is the fourth most prevalent cancer in women worldwide.^1^ It results from a chronic infection with oncogenic human papillomaviruses (HPVs),^2^ and can therefore be largely prevented through vaccination and targeted screening. Building on these opportunities, the World Health Organization (WHO) launched in 2018 its *Global strategy to accelerate the elimination of cervical cancer as a public health problem*,^3^ outlining a life-course approach combining primary, secondary, and tertiary prevention strategies.^4^ Despite this initiative, the global CC burden has increased rather than declined over the past years.^5,6^

This upward trend underscores the need for stronger public health policies to effectively address this challenge, particularly among overlooked populations such as middle-aged women (i.e. aged 35–65 years)^7^ and/or deprived women, the former falling outside vaccine deployment in many countries.^8^ Regarding middle-aged women, the few published studies to date suggest that menopausal-related discomfort during cervical cancer screening (CCS)^9^ and misconceptions about the reduced need for CC prevention after menopause due to the loss of reproductive capacity, lower sexual activity and other beliefs,^10^ constitute key perceptual and behavioural barriers to CC prevention strategies. Studies of disadvantaged women also identify low awareness, psychological barriers (e.g. fear and embarrassment), structural limitations (e.g. time constraints), sociocultural factors (e.g. inadequate family support), and religious obstacles as major contributors to lower participation in CC prevention programmes,^11–13^ particularly among specific groups such as female sex workers (FSWs).^14^

Given the knowledge gaps in CC prevention research among certain underserved groups, this study aimed to evaluate 28 possible determinants of adherence to two WHO-recommended CC prevention strategies (specifically, CCS adherence and condom use in the past year)^3^ among middle-aged women from diverse socioeconomic backgrounds. We especially focused on menopausal status and the risk of poverty and/or social exclusion (as measured by the validated European AROPE indicator)^15^ as possible understudied determinants of adherence to CC prevention strategies.

## 2. MATERIALS AND METHODS

### 2.1 Study population

This cross-sectional analytical study included middle-aged women from the INMA-Valencia (‘Environment and Childhood’)^16^ and PAPILONG (‘Human Papillomavirus in women from NGOs’)^17^ cohorts. INMA-Valencia is a long-running cohort (recruited November 2003–June 2005). Data for this study came from the ninth follow-up (2019–2021), comprising women aged 37–57 years from various socioeconomic backgrounds. PAPILONG was established between June 2021 and February 2023 and included women aged 39–64 years from socioeconomically disadvantaged strata (defined as those at risk of poverty and/or social exclusion according to the European AROPE indicator^15^ including migrants, homeless women, food-bank users, and FSWs. The inclusion criteria for both cohorts were an age range of 35–65 years and no history of hysterectomy. The final sample included 379 women (258 from INMA-Valencia and 121 from PAPILONG) living in the city of Valencia and its surrounding areas (see Figure 1).

**Figure 1.**
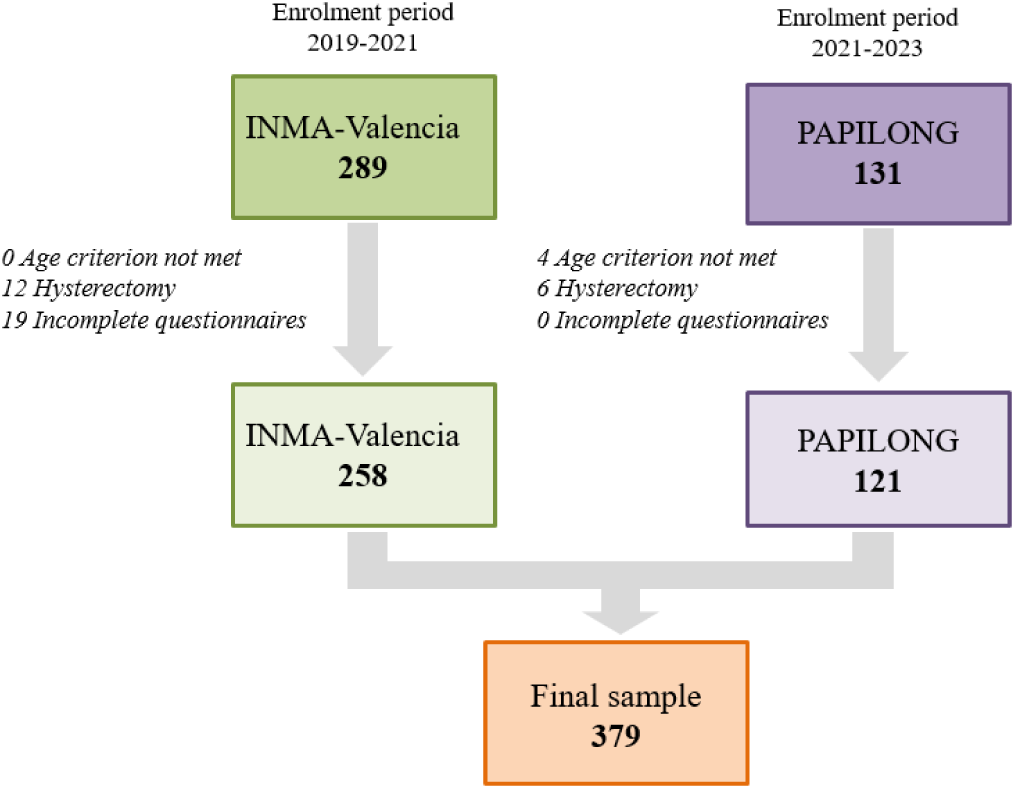
Flowchart of the study population (Valencia, Spain, 2019–2023)

### 2.2 Use of cervical cancer prevention strategies

Adherence to two WHO-recommended CC prevention strategies was assessed.^3^ First, adherence to Spanish CCS recommendations^18^ for middle-aged women was defined as having undergone cervical cytology within the previous five years (adherent vs non-adherent). For women in the PAPILONG cohort only, as the relevance of this information only emerged after completion of the INMA-Valencia follow-up, we also collected the reasons for not following CCS recommendations. These responses were categorised according to predefined themes: ‘lack of information or misperceptions’, ‘personal and emotional barriers’, and ‘structural or contextual barriers’. Second, regular condom use during vaginal intercourse in the year prior to the survey was assessed (yes/no). Finally, HPV vaccination status was recorded but it was not analysed due to the extremely low vaccination rate in our population (3.43%).

### 2.3 Studied variables

Trained interviewers administered identical structured, face-to-face questionnaires to both cohorts, consisting of multiple-choice response questions, to collect self-reported data on 28 variables: six socioeconomic indicators, ten sexual and reproductive health variables, five medical history variables, five current health and anthropometric indicators, and two lifestyle factors (see Supplementary Methods S1 and Table S1 for details).

### 2.4 Statistical analysis

Descriptive statistics are presented as frequencies (percentages) for categorical variables and as medians (25^th^ and 75^th^ percentiles [P]) for continuous variables. To identify the determinants of CCS adherence and condom use in the past year, multivariate logistic regression models were fitted separately for the full sample (n=379) and for FSWs (n=46), given the specific vulnerabilities of the latter in accessing healthcare.^14^ A sensitivity analysis was also conducted that excluded FSWs from the full sample model. To study the associated factors, a two-step procedure was applied. First, to mitigate potential overfitting, univariate logistic models were implemented for each outcome and possible candidate determinant (see Supplementary Methods S1 and Tables S1-S2). Only variables with a p-value < 0.20 in the likelihood-ratio test were retained for the multivariate models. Second, multivariate models were constructed following a backward elimination procedure, retaining variables with a p-value < 0.10 in the likelihood test. This resulted in different core models for each response variable. Menopausal status and AROPE^15^ were included as fixed covariates in the models, as evaluating their potential influence on adherence to CC prevention strategies was a key study objective. For the FSW subpopulation, menopausal status and AROPE could not be forced into the models due to the small sample size. We assessed the outliers (studentised residuals ≥ 4) and highly influential observations (Cook’s distance > 0.5). Coefficients were expressed as odds ratios (ORs) alongside their 95% confidence intervals (CIs). All statistical analyses were performed using R software (version 4.3.2; R Core Team, 2025), and associations were considered statistically significant (p-value < 0.05) or marginally significant (p-value < 0.10). Plots were generated using the ggplot2 R package.^19^

## 3. RESULTS

### 3.1 Study population

The median (P25^th^, P75^th^) age of the participants was 48.00 years (45.00, 51.00), 44.84% were AROPE (n=165), and 29.10% were postmenopausal (n=110). For FSWs (n=46), the median (P25^th^, P75^th^) age was 51.00 years (46.25, 54.00), 95.65% were AROPE (n=44), and 54.35% were postmenopausal (n=25). The median (P25^th^, P75^th^) onset age of prostitution and the duration of sex work were 37.00 years (26.75, 44.25) and 60.00 months (21.00, 135.00), respectively (see Table 1 and Supplementary Table S2).

**Table 1.**
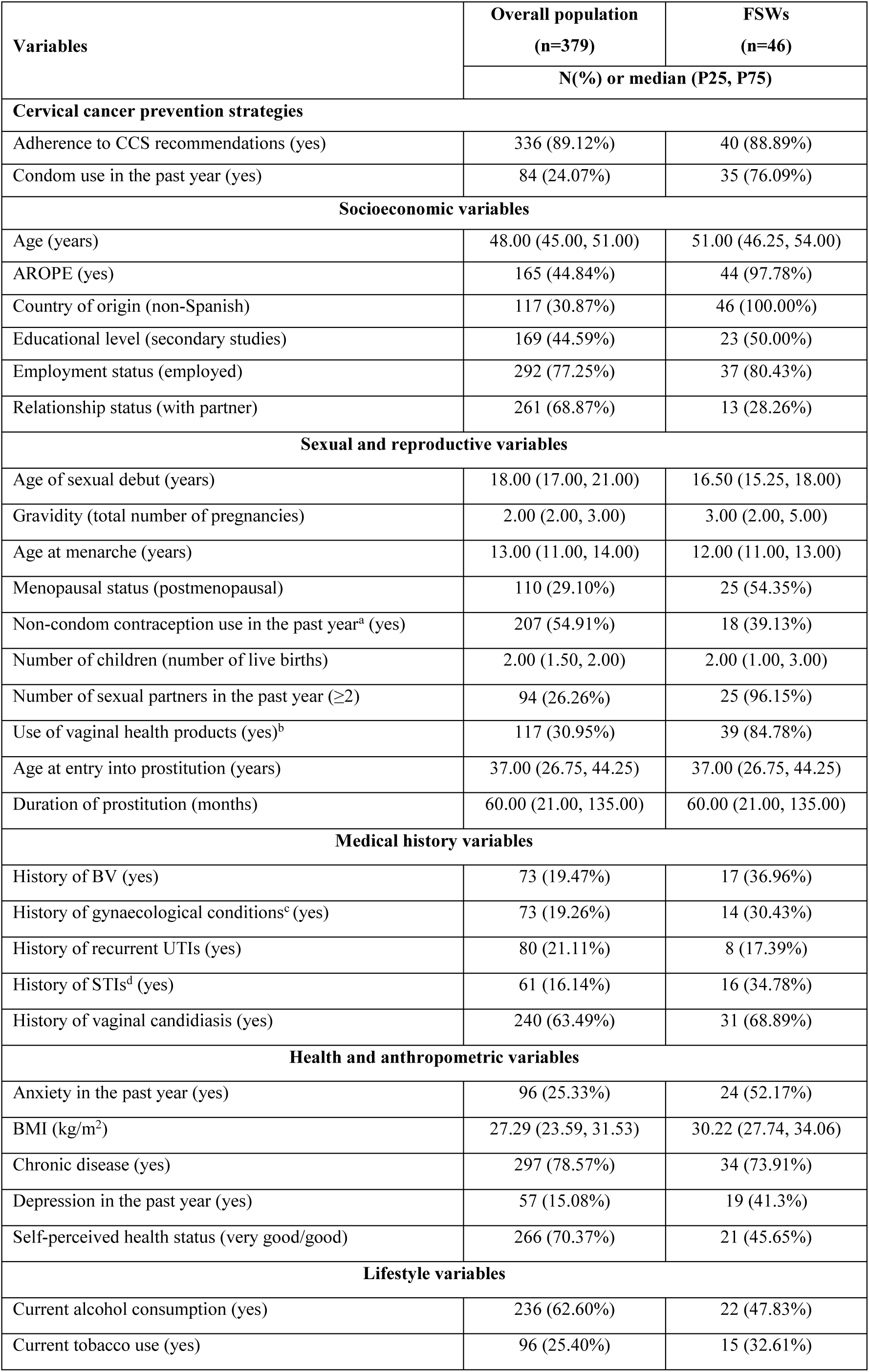
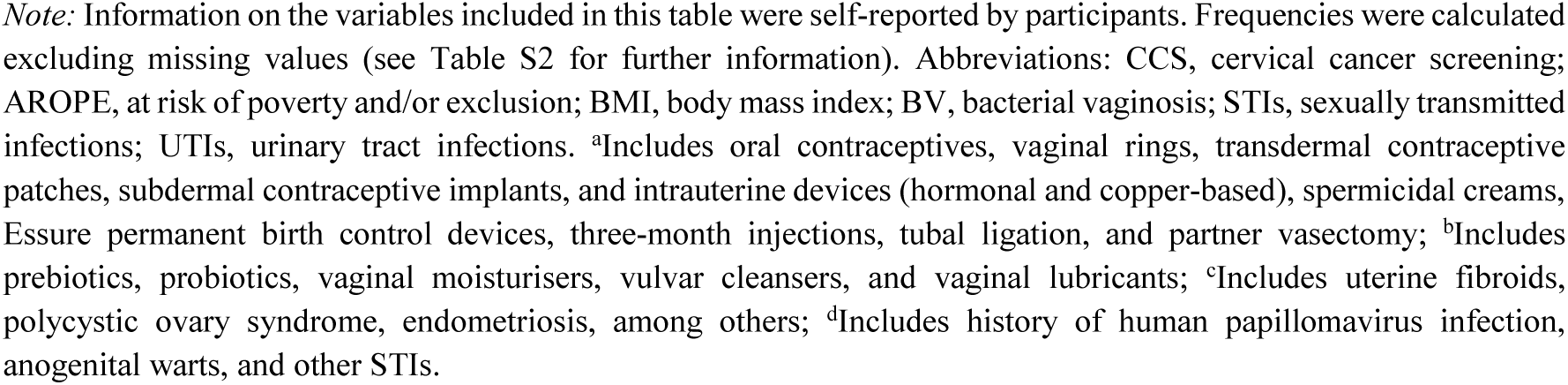
Characteristics of the study population, 2019–2023 (Valencia, Spain)

| Variables | Overall population<br>(n=379) | FSWs<br>(n=46) |
| --- | --- | --- |
|  | N(%) or median (P25, P75) |  |
| Cervical cancer prevention strategies |  |  |
| Adherence to CCS recommendations (yes) | 336 (89.12%) | 40 (88.89%) |
| Condom use in the past year (yes) | 84 (24.07%) | 35 (76.09%) |
| Socioeconomic variables |  |  |
| Age (years) | 48.00 (45.00, 51.00) | 51.00 (46.25, 54.00) |
| AROPE (yes) | 165 (44.84%) | 44 (97.78%) |
| Country of origin (non-Spanish) | 117 (30.87%) | 46 (100.00%) |
| Educational level (secondary studies) | 169 (44.59%) | 23 (50.00%) |
| Employment status (employed) | 292 (77.25%) | 37 (80.43%) |
| Relationship status (with partner) | 261 (68.87%) | 13 (28.26%) |
| Sexual and reproductive variables |  |  |
| Age of sexual debut (years) | 18.00 (17.00, 21.00) | 16.50 (15.25, 18.00) |
| Gravidity (total number of pregnancies) | 2.00 (2.00, 3.00) | 3.00 (2.00, 5.00) |
| Age at menarche (years) | 13.00 (11.00, 14.00) | 12.00 (11.00, 13.00) |
| Menopausal status (postmenopausal) | 110 (29.10%) | 25 (54.35%) |
| Non-condom contraception use in the past year<sup>a</sup> (yes) | 207 (54.91%) | 18 (39.13%) |
| Number of children (number of live births) | 2.00 (1.50, 2.00) | 2.00 (1.00, 3.00) |
| Number of sexual partners in the past year (≥2) | 94 (26.26%) | 25 (96.15%) |
| Use of vaginal health products (yes)<sup>b</sup> | 117 (30.95%) | 39 (84.78%) |
| Age at entry into prostitution (years) | 37.00 (26.75, 44.25) | 37.00 (26.75, 44.25) |
| Duration of prostitution (months) | 60.00 (21.00, 135.00) | 60.00 (21.00, 135.00) |
| Medical history variables |  |  |
| History of BV (yes) | 73 (19.47%) | 17 (36.96%) |
| History of gynaecological conditions<sup>c</sup> (yes) | 73 (19.26%) | 14 (30.43%) |
| History of recurrent UTIs (yes) | 80 (21.11%) | 8 (17.39%) |
| History of STIs<sup>d</sup> (yes) | 61 (16.14%) | 16 (34.78%) |
| History of vaginal candidiasis (yes) | 240 (63.49%) | 31 (68.89%) |
| Health and anthropometric variables |  |  |
| Anxiety in the past year (yes) | 96 (25.33%) | 24 (52.17%) |
| BMI (kg/m<sup>2</sup>) | 27.29 (23.59, 31.53) | 30.22 (27.74, 34.06) |
| Chronic disease (yes) | 297 (78.57%) | 34 (73.91%) |
| Depression in the past year (yes) | 57 (15.08%) | 19 (41.3%) |
| Self-perceived health status (very good/good) | 266 (70.37%) | 21 (45.65%) |
| Lifestyle variables |  |  |
| Current alcohol consumption (yes) | 236 (62.60%) | 22 (47.83%) |
| Current tobacco use (yes) | 96 (25.40%) | 15 (32.61%) |

*Note:* Information on the variables included in this table were self-reported by participants. Frequencies were calculated excluding missing values (see Table S2 for further information). Abbreviations: CCS, cervical cancer screening; AROPE, at risk of poverty and/or exclusion; BMI, body mass index; BV, bacterial vaginosis; STIs, sexually transmitted infections; UTIs, urinary tract infections. <sup>a</sup>Includes oral contraceptives, vaginal rings, transdermal contraceptive patches, subdermal contraceptive implants, and intrauterine devices (hormonal and copper-based), spermicidal creams, Essure permanent birth control devices, three-month injections, tubal ligation, and partner vasectomy; <sup>b</sup>Includes prebiotics, probiotics, vaginal moisturisers, vulvar cleansers, and vaginal lubricants; <sup>c</sup>Includes uterine fibroids, polycystic ovary syndrome, endometriosis, among others; <sup>d</sup>Includes history of human papillomavirus infection, anogenital warts, and other STIs.

### 3.2 Adherence to cervical cancer screening recommendations

Adherence to CCS recommendations was 89.12% (n=336) and 88.89% (n=40) for the entire population and for FSWs, respectively (see Table 1 and Supplementary Table S2). In the PAPILONG cohort, the most common reason for non-adherence was ‘lack of information or misperceptions’ (40.40%) followed by ‘personal and emotional barriers’ and ‘structural or contextual barriers’ in the same proportion (30.30%). For FSWs, ‘lack of information or misperceptions’ was also the main reason (50.00%) (see Supplementary Table S3).

In the full-sample model, middle-aged women with higher educational level had statistically significant increased odds of adhering to CCS recommendations (OR[CI95%]; 2.83[1.19–6.91] and 3.41[1.22–10.18] for secondary and university studies, respectively). Other determinants showing positive but marginally significant associations included a history of vaginal candidiasis (1.88[0.91–3.94]), the use of any non-condom contraceptive method in the past year (1.94[0.91–4.21]), a higher number of children (1.57[0.97–2.65]), and more sexual partners in the past year (2.63[0.96–8.62]) (see Figure 2 and Supplementary Table S4). A sensitivity analysis excluding FSWs yielded broadly similar results to those from the model including all participants. However, in this analysis, educational level and non-condom contraception use in the past year lost relevance, whereas AROPE became statistically significantly associated with reduced CCS adherence (OR = 1.94; CI95%: 0.89–4.18) (see Supplementary Table S4).

**Figure 2.**
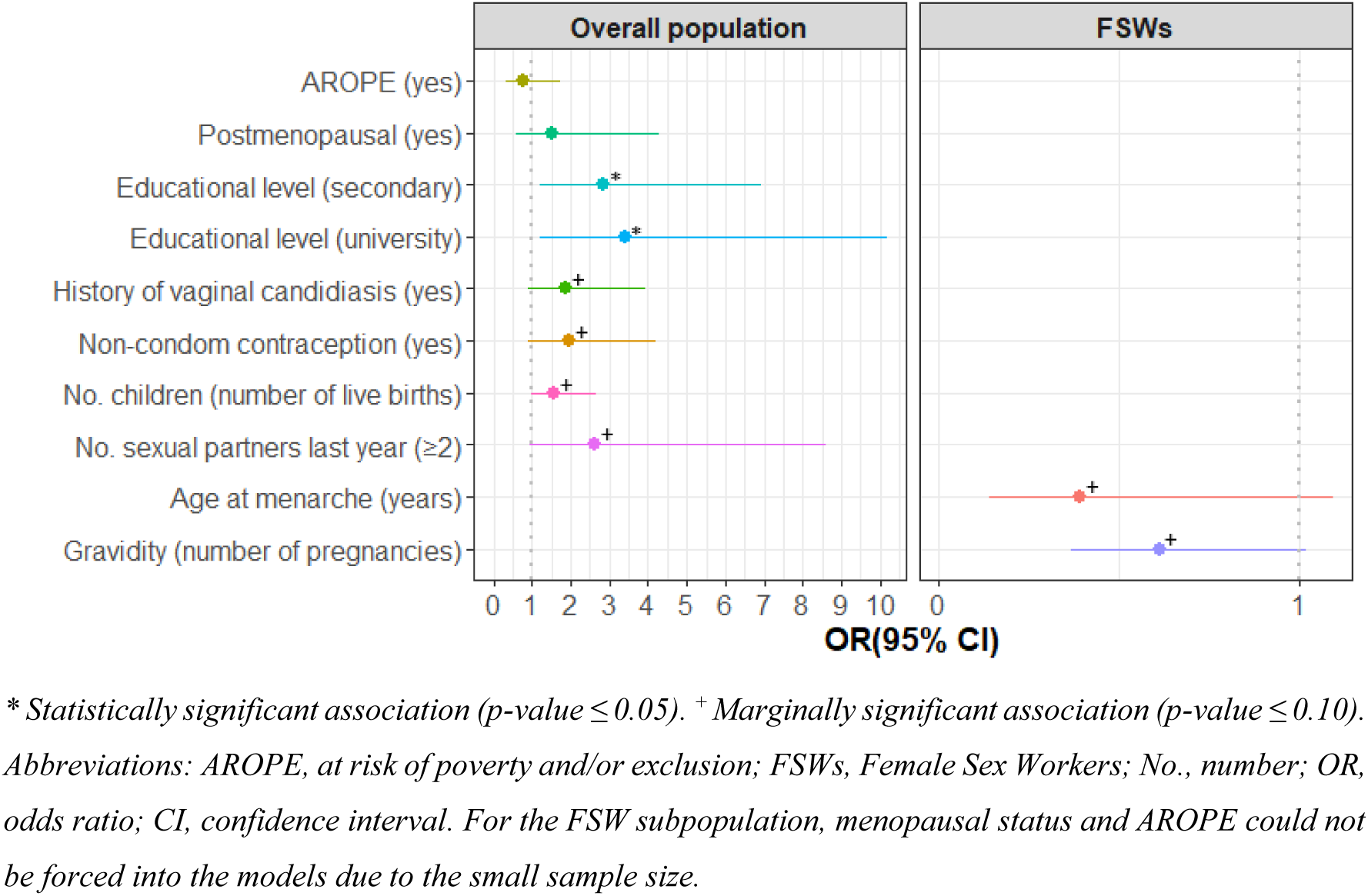
Factors associated with adherence to cervical cancer screening recommendations among middle-aged women of differing socioeconomic status (2019–2023), Valencia, Spain

In the FSW-only model, lower gravidity (0.61[0.37–1.02]]) and younger menarche age (0.39[0.14–1.09]) were marginally significantly associated with a higher likelihood of CCS adherence (see Figure 2 and Supplementary Table S4).

### 3.3 Condom use in the past year

The prevalence of condom use in the past year reported by the participants was 24.07% (n=84) and 76.09% (n=35) for the entire population and for FSWs, respectively (see Table 1 and Supplementary Table S2).

Among all participants, non-Spanish origin (2.83[1.29–6.25]), being single (2.12[1.13–3.97]), and the use of vaginal health products (2.99[1.54–5.87]) were significantly associated with a higher likelihood of condom use in the past year. In contrast, being postmenopausal (0.27[0.12–0.54]), current alcohol consumption (0.47[0.25–0.86]), and a history of recurrent urinary tract infections (UTIs) (0.41[0.18–0.87]), were associated with a lower probability of condom use. Marginally significant associations were observed for absence of chronic disease (0.53[0.27–1.05]) and sexually transmitted infection (STI) history (1.99[0.98–4.03]), both linked to condom use (see Figure 3 and Supplementary Table S5). Sensitivity analysis excluding FSWs yielded similar results to those of the main analysis; however, country of origin, UTIs history, and relationship status were no longer relevant (see Supplementary Table S5).

**Figure 3.**
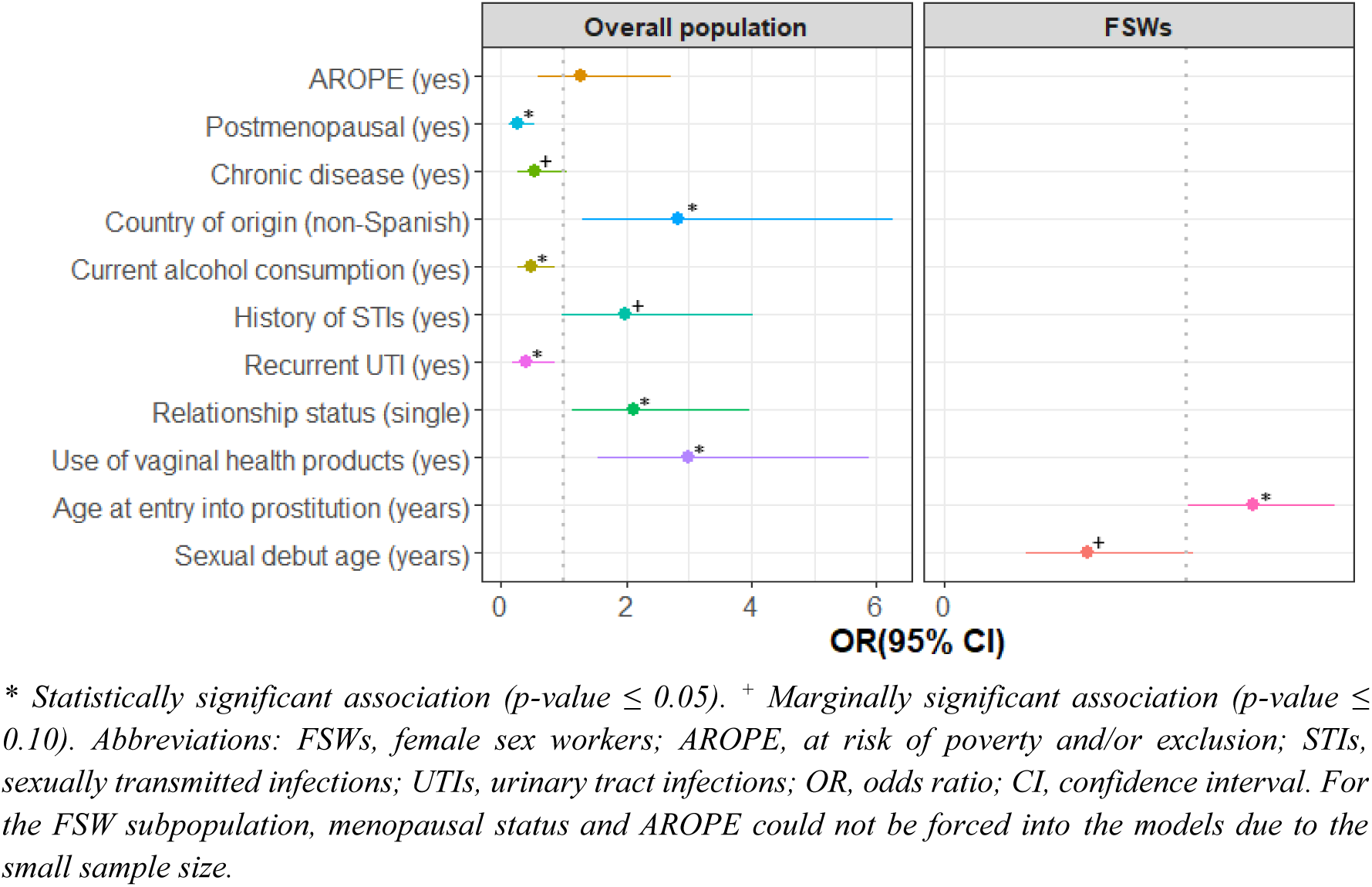
Factors associated with condom use in the past year among middle-aged women of differing socioeconomic status (2019–2023), Valencia, Spain

In the FSW-only model, older age at entry into prostitution (1.27[1.00-1.61]) was significantly associated with a higher likelihood of condom use in the past year. Furthermore, earlier sexual debut age was marginally significantly associated with condom use (0.59[0.34-1.03]) (see Figure 3 and Supplementary Table S5).

## 4. DISCUSSION

This study provides novel insights into the prevalence and determinants of CCS adherence and condom use in the past year among middle-aged women from diverse socioeconomic backgrounds (age range: 37–64 years), a population that has received limited attention in previous research. To this respect, existing literature on CCS adherence (21 studies) and condom use (12 studies), published between January 2015 and December 2025, and involving samples of between 283 and 1,610,875 women across Africa (n=7), the Americas (n=8), Asia (n=7), and Europe (n=11), generally includes much broader age ranges (15–85 years) (see Tables 2–3 and Supplementary Figures S1–S2). By focusing exclusively on midlife women, the present study addresses an important gap in the literature.

**Table 2.**
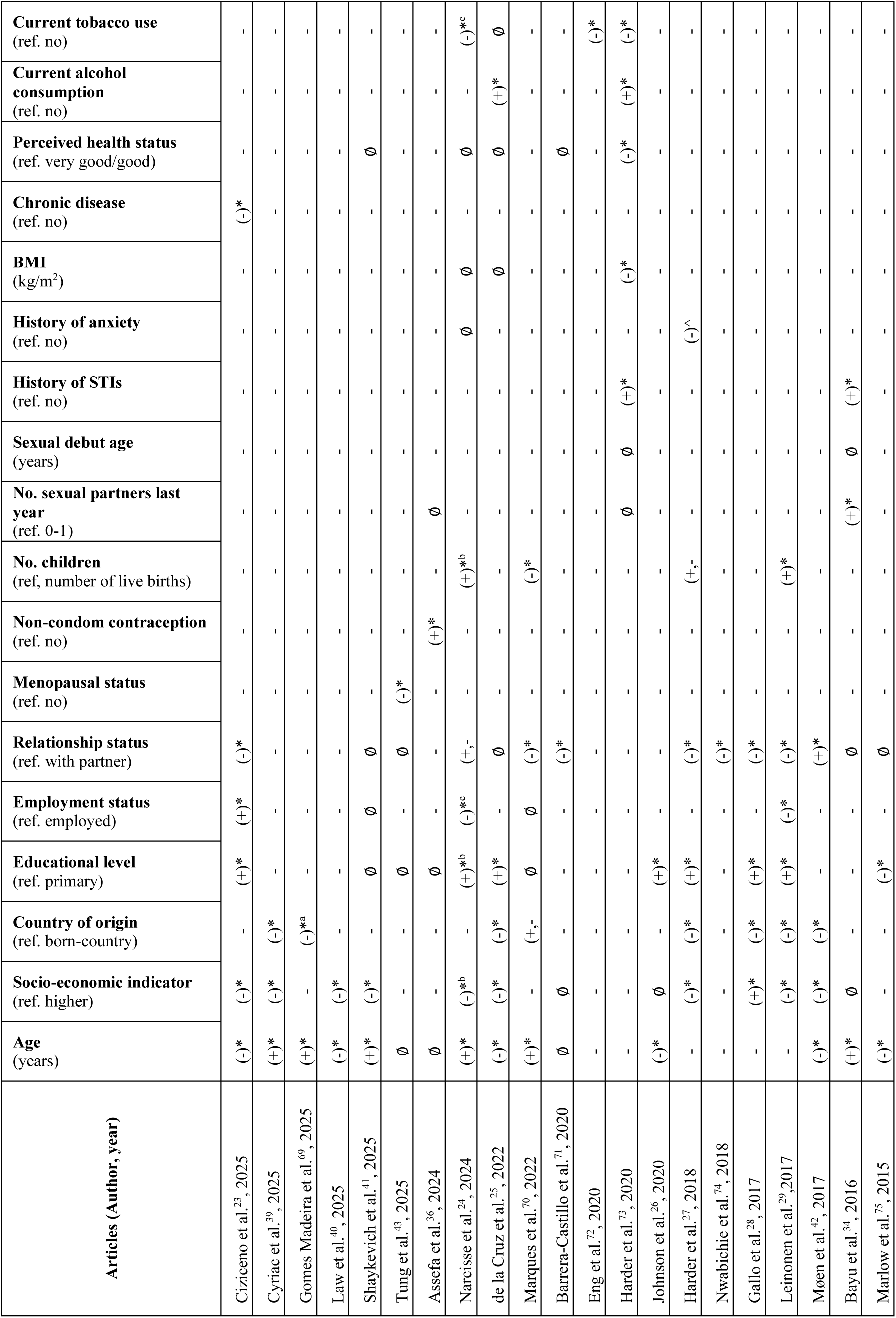
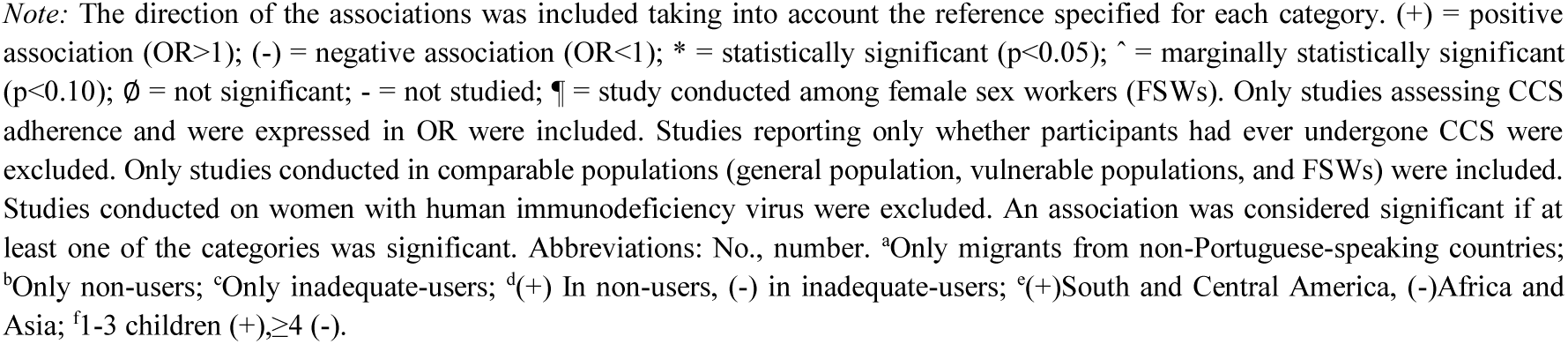
Summary of studies on determinants of cervical cancer screening adherence (January 2015–December 2025)

| Articles (Author, year) | Age (years) | Socio-economic indicator (ref. higher) | Country of origin (ref. born-country) | Educational level (ref. primary) | Employment status (ref. employed) | Relationship status (ref. with partner) | Menopausal status (ref. no) | Non-condom contraception (ref. no) | No. children (ref, number of live births) | No. sexual partners last year (ref. 0-1) | Sexual debut age (years) | History of STIs (ref. no) | History of anxiety (ref. no) | BMI (kg/m<sup>2</sup>) | Chronic disease (ref. no) | Perceived health status (ref. very good/good) | Current alcohol consumption (ref. no) | Current tobacco use (ref. no) |
| --- | --- | --- | --- | --- | --- | --- | --- | --- | --- | --- | --- | --- | --- | --- | --- | --- | --- | --- |
| Ciziceno et al.<sup>23</sup>, 2025 | (-)* | (-)* | - | (+)* | (+)* | (-)* | - | - | - | - | - | - | - | - | (-)* | - | - | - |
| Cyriac et al.<sup>39</sup>, 2025 | (+)* | (-)* | (-)* | - | - | - | - | - | - | - | - | - | - | - | - | - | - | - |
| Gomes Madeira et al.<sup>69</sup>, 2025 | (+)* | - | (-)*<sup>a</sup> | - | - | - | - | - | - | - | - | - | - | - | - | - | - | - |
| Law et al.<sup>40</sup>, 2025 | (-)* | (-)* | - | - | - | - | - | - | - | - | - | - | - | - | - | - | - | - |
| Shaykevich et al.<sup>41</sup>, 2025 | (+)* | (-)* | - | Ø | Ø | Ø | - | - | - | - | - | - | - | - | - | Ø | - | - |
| Tung et al.<sup>43</sup>, 2025 | Ø | - | - | Ø | - | Ø | (-)* | - | - | - | - | - | - | - | - | - | - | - |
| Assefa et al.<sup>36</sup>, 2024 | Ø | - | - | Ø | - | - | - | (+)* | - | Ø | - | - | - | - | - | - | - | - |
| Narcisse et al.<sup>24</sup>, 2024 | (+)* | (-)*<sup>b</sup> | - | (+)*<sup>b</sup> | (-)*<sup>c</sup> | (+,-) | - | - | (+)*<sup>b</sup> | - | - | - | Ø | Ø | - | Ø | - | (-)*<sup>c</sup> |
| de la Cruz et al.<sup>25</sup>, 2022 | (-)* | (-)* | (-)* | (+)* | - | Ø | - | - | - | - | - | - | - | Ø | - | Ø | (+)* | Ø |
| Marques et al.<sup>70</sup>, 2022 | (+)* | - | (+,-) | Ø | Ø | (-)* | - | - | (-)* | - | - | - | - | - | - | - | - | - |
| Barrera-Castillo et al.<sup>71</sup>, 2020 | Ø | Ø | - | - | - | (-)* | - | - | - | - | - | - | - | - | - | Ø | - | - |
| Eng et al.<sup>72</sup>, 2020 | - | - | - | - | - | - | - | - | - | - | - | - | - | - | - | - | - | (-)* |
| Harder et al.<sup>73</sup>, 2020 | - | - | - | - | - | - | - | - | - | Ø | Ø | (+)* | - | (-)* | - | (-)* | (+)* | (-)* |
| Johnson et al.<sup>26</sup>, 2020 | (-)* | Ø | - | (+)* | - | - | - | - | - | - | - | - | - | - | - | - | - | - |
| Harder et al.<sup>27</sup>, 2018 | - | (-)* | (-)* | (+)* | - | (-)* | - | - | (+,-) | - | - | - | (-)^ | - | - | - | - | - |
| Nwabichie et al.<sup>74</sup>, 2018 | - | - | - | - | - | (-)* | - | - | - | - | - | - | - | - | - | - | - | - |
| Gallo et al.<sup>28</sup>, 2017 | - | (+)* | (-)* | (+)* | - | (-)* | - | - | - | - | - | - | - | - | - | - | - | - |
| Leinonen et al.<sup>29</sup>, 2017 | - | (-)* | (-)* | (+)* | (-)* | (-)* | - | - | (+)* | - | - | - | - | - | - | - | - | - |
| Møen et al.<sup>42</sup>, 2017 | (-)* | (-)* | (-)* | - | - | (+)* | - | - | - | - | - | - | - | - | - | - | - | - |
| Bayu et al.<sup>34</sup>, 2016 | (+)* | Ø | - | - | - | Ø | - | - | - | (+)* | Ø | (+)* | - | - | - | - | - | - |
| Marlow et al.<sup>75</sup>, 2015 | (-)* | - | - | (-)* | - | Ø | - | - | - | - | - | - | - | - | - | - | - | - |

*Note:* The direction of the associations was included taking into account the reference specified for each category. (+) = positive association (OR>1); (-) = negative association (OR<1); \* = statistically significant (p<0.05); ^ = marginally statistically significant (p<0.10); Ø = not significant; - = not studied; ¶ = study conducted among female sex workers (FSWs). Only studies assessing CCS adherence and were expressed in OR were included. Studies reporting only whether participants had ever undergone CCS were excluded. Only studies conducted in comparable populations (general population, vulnerable populations, and FSWs) were included. Studies conducted on women with human immunodeficiency virus were excluded. An association was considered significant if at least one of the categories was significant. Abbreviations: No., number. <sup>a</sup>Only migrants from non-Portuguese-speaking countries; <sup>b</sup>Only non-users; <sup>c</sup>Only inadequate-users; <sup>d</sup>(+) In non-users, (-) in inadequate-users; <sup>e</sup>(+)South and Central America, (-)Africa and Asia; <sup>f</sup>1-3 children (+), ≥4 (-).

**Table 3.**
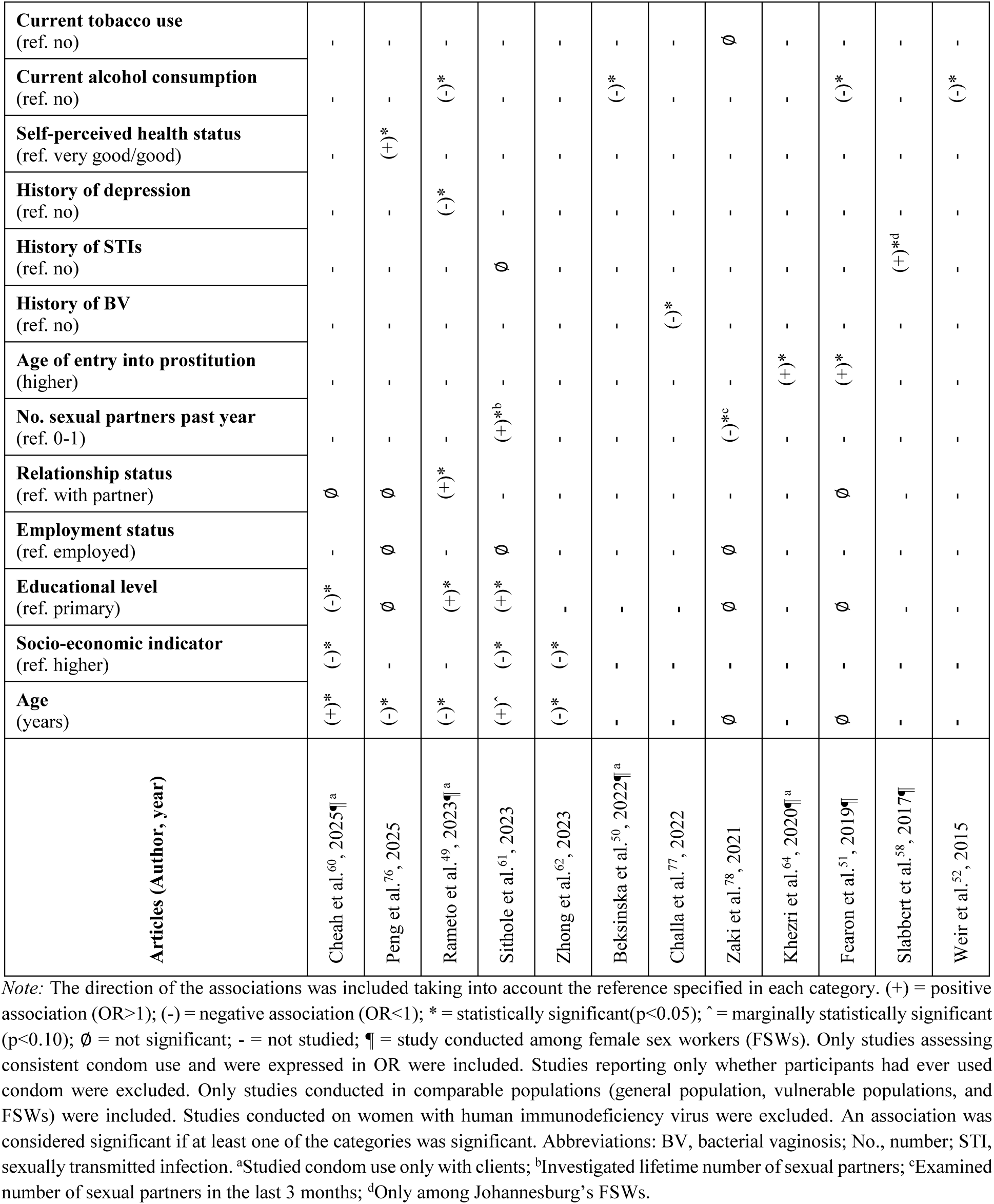
Summary of studies on determinants of condom use (January 2015 – December 2025)

### 4.1. Adherence to CCS recommendations

Adherence to CCS recommendations among our midlife population was 89.12%, slightly higher than the pooled estimates (60%–83%) reported by Zhang et al.^20^ in Western and Southern European countries with organised CCS programmes. This difference may partly reflect age-range differences between studies (34–64 years in the present study vs 18–75 years in the review),^20^ heterogeneity in CCS strategies across countries (e.g. opportunistic vs population-based),^21^ and sociocultural beliefs and norms that influence women’s participation in screening, even when such services are available.^22^

Regarding the determinants of CCS adherence in this and prior studies (see Table 2), higher education emerged as a key factor in our study, consistent with seven^23–29^ previously published papers. This association likely reflects the more limited access to information on CCS recommendations among women with lower educational levels, which may hinder informed decision-making.^30,31^ This interpretation is supported by our data on the reasons for non-adherence among the most disadvantaged participants (PAPILONG), in which ‘lack of information or misperceptions’ was the most frequently reported reason (40.40%). Similar misinformation-related barriers have been reported among underserved women^13^ and in the general population,^32^ where low health literacy has been linked to fear, embarrassment, and non-participation in CCS. Furthermore, close contact with the healthcare system also appeared to encourage women’s CCS participation in our study, albeit with weaker associations, in line with previous literature. Specifically, higher number of children was associated with greater CCS adherence in our study, consistent with two^24,29^ of the reviewed papers, possibly due to more frequent healthcare interactions during maternity-related care.^33^ In our study, a higher number of sexual partners was also linked to increased CCS adherence, in line with one prior paper,^34^ potentially reflecting greater perceived STI risk and, consequently, increased healthcare use.^35^ In addition, the use of non-condom contraceptive methods was associated with higher CCS adherence, consistent with previous findings,^36^ likely due to the more regular gynaecological visits required for the use of these contraceptives.^37^ Moreover, a history of vaginal candidiasis was likewise associated with increased CCS adherence, possibly reflecting greater healthcare-seeking behaviour, as suggested among women with vaginal candidiasis symptoms.^38^ With regard to AROPE and menopausal status, which we considered important predictors given the age range and socioeconomic diversity of our study population, only AROPE showed a marginally significant association with reduced CCS adherence in sensitivity analyses excluding FSWs, consistent with most of the prior evidence.^23–25,27,29,39–42^ No association with menopausal status was observed, although one previous study reported higher CCS participation among pre- and perimenopausal women.^43^

Given the specific vulnerabilities of FSWs in accessing healthcare^44^, we conducted a subgroup analysis focused on this population. Adherence to CCS recommendations was 22.16%, higher than the rate reported in England (12.5%),^45^ the only European country with available data included in a scoping review of this specific population group.^14^

Despite country-specific differences in CCS guidelines (population-based in England vs opportunistic in Spain at the time of the survey), which would theoretically favour higher adherence in England,^46^ this was not observed. Other factors may account for these differences, including variations in screening intervals (at the time of the survey, England recommended screening every three years for women aged 25–49 and every five years for those aged 50–64, whereas Spain recommended three-year screening for women aged 25–34 and five-year screening for those aged 35-65),^18^ as well as other unmeasured determinants. Together, these findings underscore the need for further research into this uniquely vulnerable and understudied population.

Regarding the determinants of CCS adherence among FSWs, our study found that younger age at menarche and lower gravidity were marginally associated with increased CCS adherence, with no prior literature on this topic, to our best knowledge, available for comparison. Finally, ‘lack of information or misperceptions’ was the most commonly reported barrier to CCS participation, consistent with the findings of a review of FSWs by Vimpere et al.,^14^ and likely driven by fear and embarrassment linked to limited health literacy.^13,32^

### 4.2. Condom use in the past year

Fewer than one in four participants (22.16%) reported using condoms in the 12 months prior to the survey, which is slightly lower than the proportion reported in the general Spanish population (34.10%).^47^ This difference may partially reflect age range differences (37–64 vs 35–49 years in our study and the previous Spanish paper, respectively), given that women over 50 are less likely to use condoms.^48^

In terms of the factors influencing condom use in the past year in the overall population, and compared with previous studies (Table 3), current alcohol consumption was associated with the non-use of condoms, consistent with four previous studies.^49–52^ This finding aligns with alcohol myopia theory,^53^ which suggests that alcohol abuse impairs cognitive processing, thereby increasing the likelihood of engaging in risky sexual behaviours. Lower condom use was also observed among women in stable relationships, in line with one study,^49^ and may reflect a preference by these women for alternative contraceptive methods such as oral contraceptives or long-acting reversible contraception.^54^ Additionally, non-Spanish origin was associated with higher condom use in the past year, contrasting with a previous Spanish study that did not reach statistical significance.^55^ In our sample, Spanish women were more likely to be in stable relationships than migrant women (76.7% vs 51.3%), which could favour non-condom contraceptive use and partly explain this discrepancy.^54^ Furthermore, non-menopausal women also showed a higher likelihood of condom use in the past year, consistent with findings from a study of women over 50, where reduced contraceptive need after menopause, fear of male partner response, and lower perceived risk of STIs were cited as reasons for non-use.^56^ Also, the use of condoms appeared to be driven more by perceived sexual risk than by genitourinary or chronic health conditions. In this context, women with a higher perceived risk of STIs (i.e. those with a history of STIs or vaginal product use, the latter reaching significance and probably indicative of self-care to reduce the risk of infection)^57^ reported higher rates of condom use. In contrast, participants with conditions not typically perceived as sexually transmitted (e.g. a history of recurrent UTIs or chronic disease, the former reaching statistical significance) exhibited lower condom use. While prior studies have linked STI history^58^ or certain chronic inflammatory conditions^59^ to higher condom use, no direct association has previously been reported for recurrent UTIs or vaginal product use, warranting further investigation. Finally, other examined factors, including poverty, which we had initially hypothesised to be a key determinant, were not associated with condom use in our study, despite all prior studies^60–62^ reporting lower condom use among women experiencing poverty.

Among FSWs, condom use reached 76.09%, slightly below the range reported in the review of Western European studies by Platt et al.,^63^ which ranges from approximately 82% in Spain to 100% in the Netherlands. However, given the small size of the FSWs sample (n=46), these results should be interpreted with caution. Nevertheless, the relatively high prevalence observed may indicate a higher level of health awareness among FSWs, despite nearly one quarter of our participants reporting that they did not use condoms in the past year.

Finally, in the model restricted to FSWs, an older age at entry into prostitution was significantly associated with higher condom use. This finding is consistent with two previous studies^51,64^ (see Table 3), and likely reflects differences in sexual health literacy, empowerment, and structural vulnerability, suggesting that women who enter sex work at a later age may be better equipped to negotiate condom use and maintain safer sexual practices.^64^ As for marginal associations, an earlier age at sexual debut was associated with a higher likelihood of condom use in the past year, a finding that does not align with the results of a study conducted among FSWs in Mozambique, although this association lost statistical significance in the multivariate analysis.^65^

### 4.3. Strengths and limitations

To our knowledge, this is the first study to specifically evaluate multiple CC prevention strategies among middle-aged women, a group that is often overlooked in both scientific research and public health policies related to CC and sexual health, which have traditionally focused on younger or older populations.^66,67^ By including midlife women from a range of socioeconomic backgrounds, from economically privileged individuals to disadvantaged groups such as homeless women, FSWs, and migrants, the study provides a more comprehensive and representative overview of diverse population groups. A particular strength lies in the focused analysis of FSWs, who are a key population in CC prevention efforts. This is not only because the prevalence of this neoplasm is higher in this group,^68^ but also because of the need for tailored public-health interventions that address their specific barriers to care. Adopting a life-course perspective and conducting a comprehensive assessment of multiple determinants (n = 28) enables us to identify age- and subgroup-specific factors that have largely been overlooked in previous research. The identification of reasons for CCS non-adherence within the PAPILONG cohort further advances the study by providing valuable insight into possible underlying barriers and enabling a deeper understanding of this public health issue. Another strength is our thorough literature review, although the lack of a fully systematic approach means that some relevant studies may have been overlooked. Finally, at the time of data collection, CCS in the Valencian Community was opportunistic; however, it is currently transitioning to a population-based programme (Order SCB/480/2019, of 26 April 2019). In this context, our results may help inform the development of equity-oriented CC prevention strategies for overlooked populations.

The study also has some limitations, primarily relating to sample size. In particular, the small sample size of FSWs (n=46) means that the results should be interpreted with caution. Nevertheless, given the scarcity of research focused on middle-aged FSWs, the findings presented here offer meaningful and novel insights. In addition, the low HPV vaccination prevalence (3.43%) in our study prevented us from analysing this key CC prevention outcome. This low prevalence likely reflects limited access, given that HPV vaccination is not freely available to middle-aged women in Spain and cost poses a particular barrier, especially for disadvantaged groups. Finally, as with all observational studies that rely on self-reported data, the possibility of reporting bias cannot be ruled out.

## 5. CONCLUSION

Overall, this study found high CCS adherence but low condom use in the past year among middle-aged women; in contrast, this pattern was not observed for condom use among FSWs since over three-quarters of them reported their use. The determinants of these behaviours were found to cluster across socioeconomic, sexual and reproductive health, lifestyle, and clinical domains for the whole population. Among FSWs, specific reproductive and sexual life-course characteristics were associated with both preventive behaviours. Lack of information or misperceptions emerged as the most frequently reported barrier to CCS adherence. To our knowledge, this is the first study to jointly examine the determinants of both CCS adherence and condom use exclusively among middle-aged women from diverse socioeconomic backgrounds, a population that has received little research attention. These findings could inform WHO-led CC elimination efforts and support the development of targeted prevention strategies for overlooked groups.

## Data Availability

The data supporting the findings of this study are available upon request.

## List of abbreviations

AROPE: at risk of poverty or social exclusion
CC: cervical cancer
CCS: cervical cancer screening
CIs: confidence intervals
FSWs: female sex workers
HPVs: human papillomaviruses
INMA-Valencia: Environment and Childhood
ORs: odds ratios
PAPILONG: Human Papillomavirus in women from NGOs
P: percentiles
STI: sexually transmitted infection
UTIs: urinary tract infections
WHO: World Health Organization.

## ACKNOWLEDGEMENTS

The authors would particularly like to thank all the participants for their generous collaboration. We are also grateful to the INMA-Valencia and PAPILONG researchers for their efforts and contributions to science.

## DATA AVAILABILITY STATEMENT

The data supporting the findings of this study are available upon request.

## DISCLAIMER

The authors have nothing to declare.

## ETHICS STATEMENT

The study protocol was approved by the Ethics Committee for Research of the General Directorate of Public Health and the Higher Centre for Research in Public Health (reference numbers: 20190301/09, 20190301/09/2, 20210604/10/02). Written informed consent was obtained from all participants.

## REFERENCES

1. Wu J, Jin Q, Zhang Y, Ji Y, Li J, Liu X, et al. Global burden of cervical cancer: current estimates, temporal trend and future projections based on the GLOBOCAN 2022. J Natl Cancer Cent. 2025 Jun;5(3):322–9. doi: 10.1016/j.jncc.2024.11.006

2. Walboomers JMM, Jacobs M V., Manos MM, Bosch FX, Kummer JA, Shah K V., et al. Human papillomavirus is a necessary cause of invasive cervical cancer worldwide. J Pathol. 1999 Sep;189(1):12–9. doi: 10.1002/(SICI)1096-9896(199909)189:1<12::AID-PATH431>3.0.CO;2-F

3. World Health Organization. Global strategy to accelerate the elimination of cervical cancer as a public health problem. World Health Organization. 2020. 1–56 p. https://www.who.int/publications/i/item/9789240014107

4. Simms KT, Steinberg J, Caruana M, Smith MA, Lew J Bin, Soerjomataram I, et al. Impact of scaled up human papillomavirus vaccination and cervical screening and the potential for global elimination of cervical cancer in 181 countries, 2020–99: a modelling study. Lancet Oncol. 2019;20(3):394–407. doi: 10.1016/S1470-2045(18)30836-2

5. Bray F, Laversanne M, Sung H, Ferlay J, Siegel RL, Soerjomataram I, et al. Global cancer statistics 2022: GLOBOCAN estimates of incidence and mortality worldwide for 36 cancers in 185 countries. CA Cancer J Clin. 2024 May 4;74(3):229–63. doi: https://acsjournals.onlinelibrary.wiley.com/doi/10.3322/caac.21834

6. Sung H, Ferlay J, Siegel RL, Laversanne M, Soerjomataram I, Jemal A, et al. Global Cancer Statistics 2020: GLOBOCAN Estimates of Incidence and Mortality Worldwide for 36 Cancers in 185 Countries. CA Cancer J Clin. 2021;71(3):209–49. doi: 10.3322/caac.21660

7. Harlow SD, Derby CA. Women’s Midlife Health: Why the Midlife Matters. Women’s Midlife Heal. 2015 Dec 11;1(1):5. doi: 10.1186/s40695-015-0006-7

8. Spanish Goberment M of H. Actualización de las recomendaciones de vacunación frente a VPH. Revisión de la estrategia de una dosis [Update of HPV vaccination recommendations: review of the single-dose strategy]. Madrid, Spain; 2024. https://www.sanidad.gob.es/areas/promocionPrevencion/vacunaciones/comoTrabajamos/docs/VPH_recomendaciones_vacunacion_estrategia1dosis.pdf

9. Hope KA, Moss E, Redman CWE, Sherman SM. Psycho-social influences upon older women’s decision to attend cervical screening: A review of current evidence. Prev Med (Baltim). 2017 Aug;101:60–6. doi: 10.1016/j.ypmed.2017.05.002

10. Chan C, Choi K, Wong R, Chow K, So W, Leung D, et al. Examining the Cervical Screening Behaviour of Women Aged 50 or above and Its Predicting Factors: A Population-Based Survey. Int J Environ Res Public Health. 2016 Dec 2;13(12):1195. doi: https://www.mdpi.com/1660-4601/13/12/1195

11. Bozhar H, McKee M, Spadea T, Veerus P, Heinävaara S, Anttila A, et al. Socio-economic inequality of utilization of cancer testing in Europe: A cross-sectional study. Prev Med Reports. 2022 Apr 1;26. doi: 10.1016/j.pmedr.2022.101733

12. Decker MR, Crago AL, Chu SKH, Sherman SG, Seshu MS, Buthelezi K, et al. Human rights violations against sex workers: Burden and effect on HIV. Vol. 385, The Lancet. Lancet Publishing Group; 2015. p. 186–99. doi: 10.1016/S0140-6736(14)60800-X

13. Farajimakin O. Barriers to Cervical Cancer Screening: A Systematic Review. Cureus. 2024 Jul 28;16(7). doi: 10.7759/cureus.65555

14. Vimpere L, Sami J, Jeannot E. Cervical cancer screening programs for female sex workers: a scoping review. Front Public Heal. 2023 Sep 29;11. doi: 10.3389/fpubh.2023.1226779

15. AROPE. eurostat. 2019. Glossary: At risk of poverty or social exclusion (AROPE). https://ec.europa.eu/eurostat/statistics-explained/index.php/Glossary:At_risk_of_poverty_or_social_exclusion_(AROPE)

16. Guxens M, Ballester F, Espada M, Fernández MF, Grimalt JO, Ibarluzea J, et al. Cohort Profile: the INMA--INfancia y Medio Ambiente--(Environment and Childhood) Project. Int J Epidemiol. 2012 Aug;41(4):930–40. doi: 10.1093/ije/dyr054

17. Abumallouh R, Marin N, Beneyto R, Montagud M, Nieto M, Castillejo N, et al. Cohort profile: a prospective cohort of disadvantaged women over 40 years of age in Spain (PAPILONG). Gac Sanit. 2023;37(1):S245.

18. Gobierno de España M de S. Documento de consenso sobre el programa de cribado de cáncer de cérvix en el SNS [Consensus document on the cervical cancer screening programme in the National Health System (SNS)]. Madrid, Spain; 2016. https://www.sanidad.gob.es/areas/promocionPrevencion/cribado/cribadoCancer/cancerCervix/docs/DocumentoconsensocribadoCervix.pdf

19. Wickham H. ggplot2: Elegant graphics for data analysis. 2nd ed. New York, USA: Springer-Verlag New York; 2016. doi: 10.1007/978-3-319-24277-4

20. Zhang W, Gao K, Fowkes FJI, Adeloye D, Rudan I, Song P, et al. Associated factors and global adherence of cervical cancer screening in 2019: a systematic analysis and modelling study. Global Health. 2022 Dec 9;18(1):101. doi: 10.1186/s12992-022-00890-w

21. Williams JH, Carter SM, Rychetnik L. ‘Organised’ cervical screening 45 years on: How consistent are organised screening practices? Eur J Cancer. 2014 Nov;50(17):3029–38. doi: 10.1016/j.ejca.2014.09.005

22. Sathiyaseelan G, Hashim SM, Nawi AM. Sociocultural factors influencing women’s adherence to colorectal, breast, and cervical cancer screening: a systematic review. BMC Public Health. 2025 Jun 2;25(1):2034. doi: 10.1186/s12889-025-23118-z

23. Ciziceno M, Bertolazzi A, Quaglia V. Examining cervical cancer screening adherence: how does healthism influence participation? BMC Public Health. 2025 Oct 9;25(1):3420. doi: 10.1186/s12889-025-24631-x

24. Narcisse MR, McElfish PA, Hallgren E, Pierre-Joseph N, Felix HC. Non-use and inadequate use of cervical cancer screening among a representative sample of women in the United States. Front Public Heal. 2024;12. doi: 10.3389/fpubh.2024.1321253

25. de la Cruz SP, Cebrino J. Trends and Determinants in Uptake of Cervical Cancer Screening in Spain: An Analysis of National Surveys from 2017 and 2020. Cancers (Basel). 2022;14(10). doi: . 10.3390/cancers14102481

26. Johnson NL, Head KJ, Scott SF, Zimet GD. Persistent Disparities in Cervical Cancer Screening Uptake: Knowledge and Sociodemographic Determinants of Papanicolaou and Human Papillomavirus Testing Among Women in the United States. Public Health Rep. 2020;135(4):483–91. doi: 10.1177/0033354920925094

27. Harder E, Juul KE, Jensen SM, Thomsen LT, Frederiksen K, Kjaer SK. Factors associated with non-participation in cervical cancer screening – A nationwide study of nearly half a million women in Denmark. Prev Med (Baltim). 2018;111(February):94–100. doi: 10.1016/j.ypmed.2018.02.035

28. Gallo F, Caprioglio A, Castagno R, Ronco G, Segnan N, Giordano L. Inequalities in cervical cancer screening utilisation and results: A comparison between Italian natives and immigrants from disadvantaged countries. Health Policy (New York). 2017;121(10):1072–8. doi: 10.1016/j.healthpol.2017.08.005

29. Leinonen MK, Campbell S, Klungsøyr O, Lönnberg S, Hansen BT, Nygård M. Personal and provider level factors influence participation to cervical cancer screening: A retrospective register-based study of 1.3 million women in Norway. Prev Med (Baltim). 2017;94:31–9. doi: 10.1016/j.ypmed.2016.11.018

30. Willems B, Cullati S, Prez V De, Jolidon V, Burton-Jeangros C, Bracke P. Cancer Screening Participation and Gender Stratification in Europe. J Health Soc Behav. 2020 Sep 19;61(3):377–95. doi: 10.1177/0022146520938708

31. Besó Delgado M, Cabanell JI, Molina-Barceló A, Llorens OZ, Trejo DS, Delgado B, et al. ¿Aceptan Las Mujeres De La Comunidad Valenciana La Auto-Toma Como Forma De Cribado De Cáncer De Cérvix? [Do women in the Valencian Community accept self-sampling for cervical cancer screening?]. Rev Esp Salud Pública. 2021;95:26–7.

32. Kim K, Han HR. Potential links between health literacy and cervical cancer screening behaviors: a systematic review. Psychooncology. 2016 Feb;25(2):122–30. doi: 10.1002/pon.3883

33. Seifu BL, Asnake AA, Belayneh T, Hailemariam T. Individual and community-level determinants of cervical cancer screening among Kenyan women: a multilevel analysis of a Kenya demographic and health surveys 2022. BMC Cancer. 2025 Jul 1;25(1):1067. doi: 10.1186/s12885-025-14474-5

34. Bayu H, Berhe Y, Mulat A, Alemu A. Cervical cancer screening service uptake and associated factors among age eligible women in Mekelle zone, Northern Ethiopia, 2015: A community based study using health belief model. PLoS One. 2016;11(3):1–13. doi: 10.1371/journal.pone.0149908

35. Tesfaw K, Kindie W, Mulatu K, Bogale EK. Utilisation of cervical cancer screening and factors associated with screening utilisation among women aged 30–49 years in Mertule Mariam Town, East Gojjam Zone, Ethiopia, in 2021: a cross-sectional survey. BMJ Open. 2022 Nov 22;12(11):e067229. doi: 10.1136/bmjopen-2022-067229

36. Assefa AA, Feleke T, G/Tsadik SA, Degela F, Zenebe A, Abera G. Utilization and associated factors of cervical cancer screening service among eligible women attending maternal health services at Adare General Hospital, Hawassa city, Southern Ethiopia. Sci Rep. 2024 Feb 2;14(1):2774. doi: 10.1038/s41598-024-52924-5

37. Mignot S, Ringa V, Vigoureux S, Zins M, Panjo H, Saulnier PJ, et al. Pap tests for cervical cancer screening test and contraception: analysis of data from the CONSTANCES cohort study. BMC Cancer. 2019 Dec 5;19(1):317. doi: 10.1186/s12885-019-5477-8

38. Tavares M do C, Nicol AF, Yokobatake ER, Melo MML, Vitoriano BF, Carvalho-Costa FA, et al. Evaluation of cytopathological screening results and risk factors of women who underwent Papanicolaou test in a maternity school in Fortaleza, Ceará, Brazil. Cytopathology. 2020 Nov 6;31(6):586–92. doi: 10.1111/cyt.12883

39. Cyriac J, Jenkins GD, Strelow BA, O’ Laughlin DJ, Stevens JN, MacLaughlin KL, et al. A cross-sectional analysis of factors associated with cervical cancer screening in a large midwest primary care setting. BMC Womens Health. 2025 Apr 26;25(1):204. doi: 10.1186/s12905-025-03741-z

40. Law J, Jessiman-Perreault G, Machado AA, Xu L, Chiang B, Yang H, et al. Individual-level characteristics and geospatial factors associated with cervical cancer screening participation in Alberta, Canada: a population-based cross-sectional study. BMC Public Health. 2025 Dec 1;25. doi: 10.1186/s12889-025-23898-4

41. Shaykevich A, Wojtowycz M. Factors Associated with Cervical Cancer Screening for Women in New York State: Analyzing the 2022 NYS BRFSS. J Women’s Heal. 2025 Jun 24; doi: 10.1089/jwh.2025.0027

42. Møen KA, Kumar B, Qureshi S, Diaz E. Differences in cervical cancer screening between immigrants and nonimmigrants in Norway: A primary healthcare register-based study. Eur J Cancer Prev. 2017;26(6):521–7. doi: 10.1097/CEJ.0000000000000311

43. Tung HJ, Schwarzschild G, Gopep N, Yeh MC. Cervical Cancer Screening After Menopause. Healthcare. 2025 May 16;13(10):1157. doi: 10.3390/healthcare13101157

44. Mokhwelepa LW, Ngwenya MW. Systematic Review on Public Health Problems and Barriers for Sex Workers. Open Public Health J. 2024;17(1):1–14. doi: 10.2174/0118749445264436231119172400

45. Grath-Lone LM, Marsh K, Hughes G, Ward H. The sexual health of female sex workers compared with other women in England: Analysis of cross-sectional data from genitourinary medicine clinics. Sex Transm Infect. 2014;90(4):344–50. doi: 10.1136/sextrans-2013-051381

46. Amicizia D, Piazza MF, Grammatico F, Lavieri R, Marchini F, Astengo M, et al. Organizational Determinants, Outcomes Related to Participation and Adherence to Cancer Public Health Screening: A Systematic Review. Cancers (Basel). 2025;17(11). doi: 10.3390/cancers17111775

47. Ruiz-Muñoz D, Pérez G. Women’s socioeconomic factors associated to the choice of contraceptive method in Spain. Gac Sanit. 2013 Jan;27(1):64–7. doi: 10.1016/j.gaceta.2012.05.009

48. Pilowsky D, Wu LT. Sexual risk behaviors and HIV risk among Americans aged 50 years or older: a review. Subst Abuse Rehabil. 2015 Apr;51. doi: 10.2147/SAR.S78808

49. Rameto MA, Abdella S, Ayalew J, Tessema M, Bulti J, Bati F, et al. Prevalence and factors associated with inconsistent condom use among female sex workers in Ethiopia: findings from the national biobehavioral survey, 2020. BMC Public Health. 2023 Dec 4;23(1):2407. doi: 10.1186/s12889-023-17253-8

50. Beksinska A, Nyariki E, Kabuti R, Kungu M, Babu H, Shah P, et al. Harmful Alcohol and Drug Use Is Associated with Syndemic Risk Factors among Female Sex Workers in Nairobi, Kenya. Int J Environ Res Public Health. 2022 Jun 14;19(12):7294. doi: 10.3390/ijerph19127294

51. Fearon E, Phillips A, Mtetwa S, Chabata ST, Mushati P, Cambiano V, et al. How Can Programs Better Support Female Sex Workers to Avoid HIV Infection in Zimbabwe? A Prevention Cascade Analysis. JAIDS J Acquir Immune Defic Syndr. 2019 May 1;81(1):24–35. doi: 10.1097/QAI.0000000000001980

52. Weir BW, Latkin CA. Alcohol, Intercourse, and Condom Use Among Women Recently Involved in the Criminal Justice System: Findings from Integrated Global-Frequency and Event-Level Methods. AIDS Behav. 2015;19(6):1048–60. doi: 10.1007/s10461-014-0857-1

53. Steele CM, Josephs RA. Alcohol myopia: Its prized and dangerous effects. Am Psychol. 1990;45(8):921–33. doi: 10.1037/0003-066X.45.8.921

54. Paul R, Huysman BC, Maddipati R, Madden T. Familiarity and acceptability of long-acting reversible contraception and contraceptive choice. Am J Obstet Gynecol. 2020 Apr;222(4):S884.e1-S884.e9. doi: 10.1016/j.ajog.2019.11.1266

55. López-Olmos J. Immigrant versus autochthonous women. Differences in sexual dysfunction, vaginal infections and cervical lesions. Clin Pract Res Gynaecol Obstet. 2013 Nov;40(6):242–52. doi: 10.1016/j.gine.2012.09.002

56. Cianelli R, Villegas N, Lawson S, Ferrer L, Kaelber L, Peragallo N, et al. Unique Factors that Place Older Hispanic Women at Risk for HIV: Intimate Partner Violence, Machismo, and Marianismo. J Assoc Nurses AIDS Care. 2013 Jul;24(4):341–54. doi: 10.1016/j.jana.2013.01.009

57. Erekson EA, Martin DK, Brousseau EC, Yip SO, Fried TR. Over-the-counter treatments and perineal hygiene in postmenopausal women. Menopause. 2014 Mar;21(3):281–5. doi: 10.1097/GME.0b013e31829a3216

58. Slabbert M, Venter F, Gay C, Roelofsen C, Lalla-Edward S, Rees H. Sexual and reproductive health outcomes among female sex workers in Johannesburg and Pretoria, South Africa: Recommendations for public health programmes. BMC Public Health. 2017 Jul 4;17(S3):442. doi: 10.1186/s12889-017-4346-0

59. Harris ML, Egan N, Forder PM, Bateson D, Loxton D. Patterns of contraceptive use through later reproductive years: A cohort study of Australian women with chronic disease. Tsima BM, editor. PLoS One. 2023 May 3;18(5):e0268872. doi: 10.1371/journal.pone.0268872

60. Cheah YK, Suleiman A, Ramly M. Socioeconomic and demographic factors associated with condom use among female sex workers in Malaysia. Malaysian J Public Heal Med. 2025;25(1):181–9.

61. Sithole MZ, Batidzirai JM, Yirga AA, Musekiwa A. Prevalence and factors associated with condom use among women aged 15-49 years in Rwanda using a survey logistic regression model: evidence from the 2019/20 Rwanda Demographic and Health Survey. Pan Afr Med J . 2023;46(121). doi: 10.11604/pamj.2023.46.121.41640

62. Zhong X, Chen S, Xiao H, Xiao X, Yu S, Shen Y, et al. Perceived HIV risk and factors associated with condom use among women aged 40 and older: A cross-sectional survey. Int J Nurs Sci. 2023 Oct;10(4):533–9. doi: 10.1016/j.ijnss.2023.09.017

63. Platt L, Jolley E, Rhodes T, Hope V, Latypov A, Reynolds L, et al. Factors mediating HIV risk among female sex workers in Europe: a systematic review and ecological analysis. BMJ Open. 2013 Jul;3(7):e002836. doi:10.1136/bmjopen-2013-002836

64. Khezri M, Shokoohi M, Mirzazadeh A, Karamouzian M, Sharifi H, Haghdoost A, et al. Early sex work initiation and its association with condomless sex and sexually transmitted infections among female sex workers in Iran. Int J STD AIDS. 2020 Jun 13;31(7):671–9. doi: 10.1177/0956462420913431

65. Muleia R, Banze AR, Damião SL, Baltazar CS. Patterns of inconsistent condom use and risky sexual behaviors among female sex workers in Mozambique. BMC Public Health. 2024 Oct 4;24(1):2711. doi: 10.1186/s12889-024-20236-y

66. James JJ, Klevenow EA, Atkinson MA, Vosters EE, Bueckers EP, Quinn ME, et al. Underrepresentation of women in exercise science and physiology research is associated with authorship gender. J Appl Physiol. 2023 Oct 1;135(4):932–42. doi: 10.1152/japplphysiol.00377.2023

67. WHO. World Health Organization. Women, Ageing and Health: A Framework for Action Focus on gender Women, Ageing and Health: A Framework for Action Focus on gender. WHO Library Cataloguing. 2007.

68. Farahmand M, Moghoofei M, Dorost A, Abbasi S, Monavari SH, Kiani SJ, et al. Prevalence and genotype distribution of genital human papillomavirus infection in female sex workers in the world: A systematic review and meta-analysis. BMC Public Health. 2020;20(1):1–14. doi: 10.1186/s12889-020-09570-z

69. Gomes Madeira C, Marotta C, Ditter AG, Battista Raviglione MC. Participation in cervical cancer screening among migrants and non-migrants in primary healthcare in Lisbon : a register-based study. BMJ Glob Heal. 2025;10(12):e019061. doi: 10.1136/bmjgh-2025-019061

70. Marques P, Geraldes M, Gama A, Heleno B, Dias S. Non-attendance in cervical cancer screening among migrant women in Portugal: A cross-sectional study. Women’s Heal. 2022 Jan 17;18:1–9. doi: 10.1177/17455057221093034

71. Barrera-Castillo M, Fernández-Peña R, del Valle-Gómez M del O, Fernández-Feito A, Lana A. Social integration and gynecologic cancer screening of immigrant women in Spain. Gac Sanit. 2020;34(5):468–73. doi: 10.1016/j.gaceta.2019.01.002

72. Eng VA, David SP, Li S, Ally MS, Stefanick M, Tang JY. The association between cigarette smoking, cancer screening, and cancer stage: a prospective study of the women’s health initiative observational cohort. BMJ Open. 2020 Aug 13;10(8):e037945. doi: 10.1136/bmjopen-2020-037945

73. Harder E, Hertzum-Larsen R, Frederiksen K, Kjær SK, Thomsen LT. Non-participation in cervical cancer screening according to health, lifestyle and sexual behavior: A population-based study of nearly 15,000 Danish women aged 23–45 years. Prev Med (Baltim). 2020;137(October 2019):106119. doi: 10.1016/j.ypmed.2020.106119

74. Nwabichie CC, Manaf RA, Ismail SB. Factors affecting uptake of cervical cancer screening among African Women in Klang Valley, Malaysia. Asian Pacific J Cancer Prev. 2018;19(3):825–31. doi: 10.22034/APJCP.2018.19.3.825

75. Marlow LA V, Wardle J, Waller J. Understanding cervical screening non-attendance among ethnic minority women in England. Br J Cancer. 2015 Sep 14;113(5):833–9. doi: 10.1038/bjc.2015.248

76. Peng X, Wang B, Lu Y, Li X, Li Y, Ouyang L, et al. PrEP-eligible behaviours and condom use among sexually active older adults in China: Findings from the sexual well-being (SWELL) study. Public Health. 2025 Apr;241(September 2024):164–70. doi: 10.1016/j.puhe.2025.02.014

77. Challa A, Kachhawa G, Sood S, Upadhyay AD, Dwivedi SN, Gupta S. Correlates of bacterial vaginosis among women from North India. Int J STD AIDS. 2022 Jun 24;33(7):666–71. doi: 10.1177/09564624221091743

78. Zaki SA, Naous J, Ghanem A, Abou Abbas D, Tomb R, Ghosn J, et al. Prevalence of STIs, sexual practices and substance use among 2083 sexually active unmarried women in Lebanon. Sci Rep. 2021;11(1):1–11. doi: 10.1038/s41598-021-89258-5

