## Supplementary Material for "Determinants of Adherence to Cervical Cancer Prevention Strategies Among Socially Diverse Middle-Aged Women"

#### Supplementary Methods

**Supplementary Methods S1.** Description of the variables studied

#### Supplementary Tables

**Supplementary Table S1.** Studied variables related to adherence to cervical cancer prevention strategies

**Supplementary Table S2.** Characteristics of the study population (2019–2023), Valencia, Spain

**Supplementary Table S3.** Barriers to adherence to cervical cancer screening recommendations among PAPILONG cohort participants (2021–2023), Valencia, Spain

**Supplementary Table S4.** Factors associated with adherence to cervical cancer screening recommendations among middle-aged women of differing socioeconomic status (2019–2023), Valencia, Spain

**Supplementary Table S5.** Factors associated with condom use in the past year among middle-aged women of differing socioeconomic status (2019–2023), Valencia, Spain

#### Supplementary Figures

**Supplementary Figure S1.** Summary of studies on the determinants of cervical cancer screening adherence (January 2015–December 2025)

**Supplementary Figure S2.** Summary of studies on the determinants of condom use (January 2015–December 2025)

### Supplementary Methods

**Supplementary Methods S1.** Description of the variables studied

We gathered information on six sociodemographic and economic indicators: age (years); AROPE (indicator of poverty or social exclusion;[1] yes/no); country of origin (Spain/non-Spanish); educational level (primary/secondary/university studies); employment status (employed/unemployed); and relationship status (single/with partner).

Ten sexual and reproductive health variables were assessed: age of sexual debut (years); non-condom contraception in the past year (yes/no); age at menarche (years); menopausal status (not postmenopausal/ postmenopausal); number of children (number of live births); number of sexual partners in the past year ($\leq 1/\geq 2$); gravidity (total number of pregnancies); use of vaginal health products (including prebiotics, probiotics, vaginal moisturizers, vulvar cleansers, and vaginal lubricants; yes/no); age at entry into prostitution (age); and duration of prostitution (months).

Five medical history variables were included: history of bacterial vaginosis (BV; yes/no); gynaecological conditions (including uterine fibroids, polycystic ovary syndrome, and endometriosis, among others; yes/no); recurrent urinary tract infections (UTIs; yes/no); sexually transmitted infections (STIs, including history of human papillomavirus [HPV] infection, anogenital warts, and other STIs; yes/no); and history of vaginal candidiasis (yes/no).

Five current health and anthropometric indicators were assessed: self-reported anxiety in the past year (yes/no); body mass index (BMI; $kg/m^2$); presence of chronic disease (yes/no); self-reported depression in the past year (yes/no); and perceived health status (very good-good/fair/bad-very bad).

Finally, two lifestyle factors were examined: current alcohol consumption (yes/no) and current tobacco use (yes/no).

### Supplementary Tables

#### **Supplementary Table S1.** Studied variables related to adherence to cervical cancer prevention strategies

| **Socioeconomic** | **Sexual and reproductive health** | **Medical history** | **Health and anthropometric** | **Lifestyle** |
|---|---|---|---|---|
| • Age<br>• AROPE<br>• Country of origin<br>• Educational level<br>• Employment status<br>• Relationship status | • Age at menarche<br>• Age of sexual debut<br>• Gravidity<br>• Menopausal status<br>• Non-condom contraception use in the past year[a]<br>• Number of children<br>• Number of sexual partners in the past year<br>• Use of vaginal health products[b]<br>• Age at entry into prostitution<br>• Duration of prostitution | *History of:*<br>• BV<br>• Gynaecological conditions[c]<br>• Recurrent UTIs<br>• STIs[d]<br>• Vaginal candidiasis | • Self-reported anxiety in the past year<br>• BMI<br>• Chronic disease<br>• Self-reported depression in the past year<br>• Perceived health status | • Current alcohol consumption<br>• Current tobacco use |

*Note:* Information on the variables included in this table was self-reported by participants. *A*bbreviations: AROPE, at risk of poverty or exclusion; *BMI*, body mass index; *BV*, bacterial vaginosis; *STIs*, sexually transmitted infections; *UTIs*, urinary tract infections. [a] Includes oral contraceptives, vaginal rings, transdermal contraceptive patches, subdermal contraceptive implants, and intrauterine devices (hormonal and copper-based), spermicidal creams, Essure permanent birth control devices, three-month injections, tubal ligation and partner vasectomy. [b] Includes prebiotics, probiotics, vaginal moisturizers, vulvar cleansers, and vaginal lubricants. [c] Includes uterine fibroids, polycystic ovary syndrome, endometriosis, among others. [d] Includes history of human papillomavirus infection, anogenital warts, and other STIs.

**Supplementary Table S2.** Characteristics of the study population (2019–2023), Valencia, Spain

| **Variables** | **Overall population (n=379)** N (%) or Median (P25, P75) | **FSWs (n=46)** N (%) or Median (P25, P75) |
|---|---|---|
| **Cervical cancer prevention strategies** | | |
| Adherence to CCS recommendations | | |
| No | 41 (10.88%) | 5 (11.11%) |
| Yes | 336 (89.12%) | 40 (88.89%) |
| Condom use in the past year | | |
| No | 265 (75.93%) | 11 (23.91%) |
| Yes | 84 (24.07%) | 35 (76.09%) |
| **Socioeconomic variables** | | |
| Age (years) | 48.00 (45.00, 51.00) | 51.00 (46.25, 54.00) |
| AROPE | | |
| No | 203 (55.16%) | 1 (2.22%) |
| Yes | 165 (44.84%) | 44 (97.78%) |
| Country of origin | | |
| Spain | 262 (69.13%) | 0 (0.00%) |
| Non-Spanish | 117 (30.87%) | 46 (100.00%) |
| Educational level | | |
| Primary studies | 92 (24.27%) | 19 (41.30%) |
| Secondary studies | 169 (44.59%) | 23 (50.00%) |
| University studies | 118 (31.13%) | 4 (8.70%) |
| Employment status | | |
| Employed | 292 (77.25%) | 37 (80.43%) |
| Unemployed | 86 (22.75%) | 9 (19.57%) |
| Relationship status | | |
| With partner | 261 (68.87%) | 13 (28.26%) |
| Single | 118 (31.13%) | 33 (71.74%) |
| **Sexual and reproductive variables** | | |
| Age at sexual debut (years) | 18.00 (17.00, 21.00) | 16.50 (15.25, 18.00) |
| Gravidity (total number of pregnancies) | 2.00 (2.00, 3.00) | 3.00 (2.00, 5.00) |
| Menarche age (years) | 13.00 (11.00, 14.00) | 12.00 (11.00, 13.00) |
| Menopausal status | | |
| Non-postmenopausal | 268 (70.90%) | 21 (45.65%) |
| Postmenopausal | 110 (29.10%) | 25 (54.35%) |
| Non-condom contraception use in the past year[a] | | |
| No | 170 (45.09%) | 28 (60.87%) |
| Yes | 207 (54.91%) | 18 (39.13%) |
| Number of children (number of live births) | 2.00 (1.50, 2.00) | 2.00 (1.00, 3.00) |

| Variables | Overall population (n=379)<br>N (%) or Median (P25, P75) | FSWs (n=46)<br>N (%) or Median (P25, P75) |
|---|---|---|
| Number of sexual partners in the past year | | |
| ≤1 | 264 (73.74%) | 1 (3.85%) |
| ≥2 | 94 (26.26%) | 25 (96.15%) |
| Use of vaginal health products[b] | | |
| No | 261 (69.05%) | 7 (15.22%) |
| Yes | 117 (30.95%) | 39 (84.78%) |
| Age at entry into prostitution (years) | 37.00 (26.75, 44.25) | 37.00 (26.75, 44.25) |
| Duration of prostitution (months) | 60.00 (21.00, 135.00) | 60.00 (21.00, 135.00) |
| **Medical history variables** | | |
| History of BV | | |
| No | 302 (80.53%) | 29 (63.04%) |
| Yes | 73 (19.47%) | 17 (36.96%) |
| History of gynaecological conditions[c] | | |
| No | 306 (80.74%) | 32 (69.57%) |
| Yes | 73 (19.26%) | 14 (30.43%) |
| History of recurrent UTIs | | |
| No | 299 (78.89%) | 38 (82.61%) |
| Yes | 80 (21.11%) | 8 (17.39%) |
| History of STIs[d] | | |
| No | 317 (83.86%) | 30 (65.22%) |
| Yes | 61 (16.14%) | 16 (34.78%) |
| History of vaginal candidiasis | | |
| No | 138 (36.51%) | 14 (31.11%) |
| Yes | 240 (63.49%) | 31 (68.89%) |
| **Health and anthropometric variables** | | |
| Self-reported anxiety in the past year | | |
| No | 283 (74.67%) | 22 (47.83%) |
| Yes | 96 (25.33%) | 24 (52.17%) |
| BMI (kg/m$^2$) | 27.29 (23.59, 31.53) | 30.22 (27.74, 34.06) |
| Chronic disease | | |
| No | 81 (21.43%) | 12 (26.09%) |
| Yes | 297 (78.57%) | 34 (73.91%) |
| Self-reported depression in the past year | | |
| No | 321 (84.92%) | 27 (58.70%) |
| Yes | 57 (15.08%) | 19 (41.30%) |
| Perceived health status | | |
| Very good/Good | 266 (70.37%) | 21 (45.65%) |
| Fair | 84 (22.22%) | 18 (39.13%) |

| Variables | Overall population (n=379)<br>N (%) or Median (P25, P75) | FSWs (n=46)<br>N (%) or Median (P25, P75) |
|---|---|---|
| Bad/Very bad | 28 (7.41%) | 7 (15.22%) |
| **Lifestyle variables** | | |
| Current alcohol consumption | | |
| No | 141 (37.40%) | 24 (52.17%) |
| Yes | 236 (62.60%) | 22 (47.83%) |
| Current tobacco use | | |
| No | 282 (74.60%) | 31 (67.39%) |
| Yes | 96 (25.40%) | 15 (32.61%) |

*Note:* Frequencies are calculated excluding missing values. All the variables included in this table were self-reported by participants. *A*bbreviations: AROPE, at risk of poverty or exclusion; BMI, body mass index; BV, bacterial vaginosis; CCS, cervical cancer screening; FSWs, female sex workers; STIs, sexually transmitted infections; UTIs, urinary tract infections. [a] Includes oral contraceptives, vaginal rings, transdermal contraceptive patches, subdermal contraceptive implants, and intrauterine devices (hormonal and copper-based), spermicidal creams, Essure permanent birth control devices, three-month injections, tubal ligation and partner vasectomy. [b] Includes prebiotics, probiotics, vaginal moisturizers, vulvar cleansers, and vaginal lubricants. [c] Includes uterine fibroids, polycystic ovary syndrome, endometriosis, among others. [d] Includes history of human papillomavirus infection, anogenital warts and other STIs.

**Supplementary Table S3.** Barriers to adherence to cervical cancer screening recommendations among PAPILONG cohort participants (2021–2023), Valencia, Spain

| | Number of times barrier listed (%) | |
|---|---|---|
| **Categorised barrier** | **PAPILONG** | **FSWs** |
| Personal and emotional barriers[a] | 30.30 | 16.67 |
| Lack of information or misperceptions[b] | 40.40 | 50.00 |
| Structural or contextual barriers[c] | 30.30 | 33.33 |

Abbreviations: FSWs female sex workers. [a]Includes fear, anxiety, lack of motivation, forgetfulness, or avoidance of medical settings. [b]Includes misinformation, incorrect beliefs, and low perceived risk. [c]Includes challenges related to country of origin, integration into a new health system, or negative experiences with screening. Qualitative reasons for non-adherence to CCS recommendations were collected only for women in the PAPILONG cohort, as the relevance of this information became evident after completion of recruitment for the INMA cohort.

**Supplementary Table S4.** Factors associated with adherence to cervical cancer screening recommendations among middle-aged women of differing socioeconomic status (2019–2023), Valencia, Spain

| | Overall population (n=379) | | FSWs (n=46) | | Sensitivity model excluding FSWs (n=333) | |
|---|---|---|---|---|---|---|
| **Variables** | OR [95% CI] | p-value | OR [95% CI] | p-value | OR [95% CI] | p-value |
| **AROPE:** yes vs no (ref) | 0.76 [0.33–1.75] | 0.510 | – | – | 0.50 [0.23–1.09] | $0.078^{+}$ |
| **Menopausal status:** postmenopausal vs non-postmenopausal (ref) | 1.52 [0.61–4.27] | 0.391 | – | – | 2.01 [0.76–6.42] | 0.191 |
| **Educational level** | | | | | | |
| $\leq$ Primary studies (ref) | Ref | Ref | – | – | – | – |
| Secondary studies | 2.83 [1.19–6.91] | $0.020^{*}$ | – | – | – | – |
| University studies | 3.41 [1.22–10.18] | $0.022^{*}$ | – | – | – | – |
| **History of vaginal candidiasis:** yes vs no (ref) | 1.88 [0.91–3.94] | $0.088^{+}$ | – | – | 2.11 [1.02–4.44] | $0.046^{*}$ |
| **Non-condom contraception in the past year**$^{a}$: yes vs no (ref) | 1.94 [0.91–4.21] | $0.088^{+}$ | – | – | – | – |
| **No. of children** (number of live births) | 1.57 [0.97–2.65] | $0.081^{+}$ | – | – | 1.93 [1.13–3.42] | $0.021^{*}$ |
| **No.of sexual partners in the past year:** $\geq$2 vs $\leq$1 (ref) | 2.63 [0.96–8.62] | $0.079^{+}$ | – | – | 3.45 [1.09–15.45] | $0.059^{+}$ |
| **Age at menarche** (years) | – | – | 0.39 [0.14–1.09] | $0.073^{+}$ | – | – |
| **Gravidity** (total number of pregnancies) | – | – | 0.61 [0.37–1.02] | $0.058^{+}$ | – | – |

*Note:* $^{*}$Statistically significant association ($p \leq 0.05$). $^{+}$ Marginally significant association ($p \leq 0.10$). –: Variable not retained in the final model. Menopausal status and AROPE were included as fixed covariates in the models, as a key objective of our study was to evaluate their potential influence on adherence to CC prevention strategies. For the FSW subpopulation, menopausal status and AROPE could not be forced into the models due to the small sample size. Sensitivity analyses: overall population excluding FSWs. *A*bbreviations: AROPE, at risk of poverty or social exclusion; CI, confidence interval; FSWs, female sex workers; No., number; OR, odds ratio. $^{a}$Includes oral contraceptives, vaginal rings, transdermal contraceptive patches, subdermal contraceptive implants, and intrauterine devices, (hormonal and copper-based), spermicidal creams, Essure permanent birth control devices, three-month injections, tubal ligation and partner vasectomy.

**Supplementary Table S5.** Factors associated with condom use in the past year among middle-aged women of differing socioeconomic status (2019–2023), Valencia, Spain

| | Overall population (n=379) | | FSWs (n=46) | | Sensitivity model excluding FSWs (n=333) | |
|---|---|---|---|---|---|---|
| **Variables** | OR [95% CI] | p-value | OR [95% CI] | p-value | OR [95% CI] | p-value |
| **AROPE:** yes vs no (ref) | 1.28 [0.59–2.72] | 0.520 | – | – | 1.38 [0.67–2.81] | 0.373 |
| **Menopausal status:** postmenopausal vs non-postmenopausal (ref) | 0.27 [0.12–0.54] | 0.000* | – | – | 0.20 [0.07–0.50] | 0.001* |
| **Chronic disease:** yes vs no (ref) | 0.53 [0.27–1.05] | 0.065+ | – | – | 0.47 [0.22–1.02] | 0.051+ |
| **Country of origin:** Spain vs non-Spanish (ref) | 2.83 [1.29–6.25] | 0.010* | – | – | – | – |
| **Current alcohol consumption:** yes vs no (ref) | 0.47 [0.25–0.86] | 0.014* | – | – | 0.37 [0.18–0.73] | 0.005* |
| **History of STIs**[a]: yes vs no (ref) | 1.99 [0.98–4.03] | 0.056+ | – | – | 2.66 [1.13–6.14] | 0.022* |
| **History of recurrent UTIs:** yes vs no (ref) | 0.41 [0.18–0.87] | 0.027* | – | – | – | – |
| **Relationship status:** single vs with partner (ref) | 2.12 [1.13–3.97] | 0.019* | – | – | – | – |
| **Use of vaginal health products**[b]: yes vs no (ref) | 2.99 [1.54–5.87] | 0.001* | – | – | 2.56 [1.21–5.41] | 0.014* |
| **Age at entry into prostitution** (years) | – | – | 1.27 [1.00–1.61] | 0.046* | – | – |
| **Sexual debut age** (years) | – | – | 0.59 [0.34–1.03] | 0.063+ | – | – |

*Note:* *Statistically significant association ($p \leq 0.05$). + Marginally significant association ($p \leq 0.10$). –: Variable not retained in the final model. Menopausal status and AROPE were included as fixed covariates in the models, as a key objective of our study was to evaluate their potential influence on adherence to CC prevention strategies. For the FSW subpopulation, menopausal status and AROPE could not be forced into the models due to the small sample size. Sensitivity analyses: overall population excluding FSWs. *A*bbreviations: AROPE, at risk of poverty and/or social exclusion; CI, confidence interval; FSWs, female sex workers; OR, odds ratio; STIs, sexually transmitted infections; UTIs, urinary tract infections. [a]Includes history of human papillomavirus infection, anogenital warts, and other STIs. [b]Includes prebiotics, probiotics, vaginal moisturizers, vulvar cleansers, and vaginal lubricants.

### Supplementary Figures

**Supplementary Figure S1.** Summary of studies on the determinants of cervical cancer screening adherence (January 2015–December 2025)

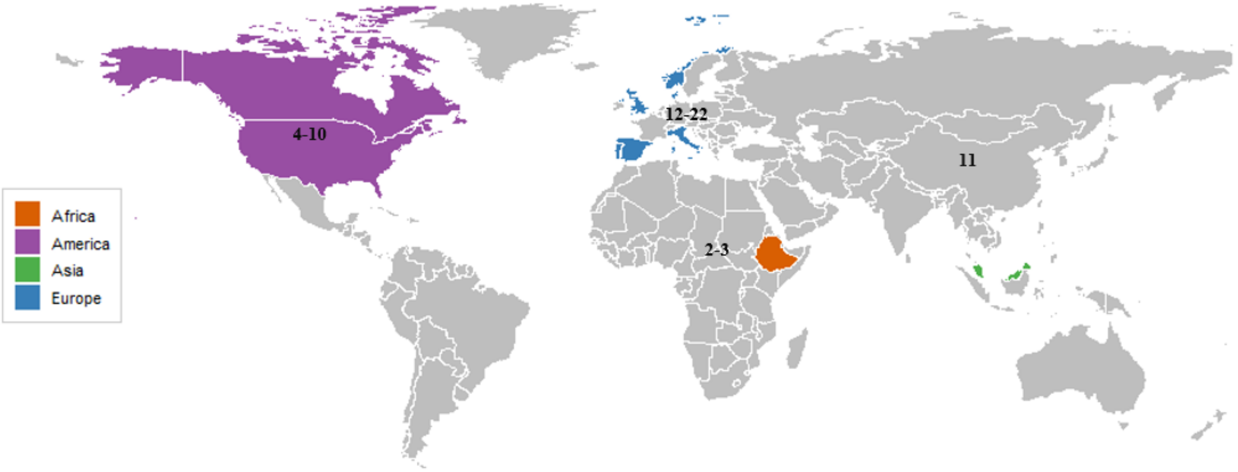


*Note:* This figure shows the countries in which at least one study on the topic was conducted between January 2015 and December 2025, using colour codes by continent. Numbers correspond to the bibliographic references listed below and include information on participants' age range and study location.

Africa:[2]Assefa et al., 2024. Age range: 30–49 years. Ethiopia;[3]Bayu et al., 2016. Age range: $\geq$21 years. Ethiopia.

America:[4]Cyriac et al., 2025. Age range: 21–65 years. USA;[5]Law et al., 2025 years. Age range: 28–69 years. USA;[6]Shaykevich et al., 2025. Age range: 30–65 years. USA;[7]Tung et al., 2025. Age range: $\geq$45 years. USA;[8]Narcisse et al., 2024. Age range: 21–65 years. USA;[9]Eng et al., 2020. Age range: postmenopausal. USA;[10]Johnson et al., 2020. Age range: 18–66 years. USA.

Asia:[11]Nwabichie et al., 2018. Age range: 18–69 years. Malaysia.

Europe:[12]Ciziceno et al., 2025. Age range: 25–64 years. Italy;[13]Gomes Madeira et al., 2025. Age range: 25–60 years. Portugal;[14]de la Cruz et al., 2022. Age range: 25–65 years. Spain;[15]Marques et al., 2022. Age range: $>$20 years. Portugal;[16]Barrera-Castillo et al., 2020. Age range: 18–75 years. Spain;[17]Harder et al., 2020. Age range: 23–45 years. Denmark;[18]Harder et al., 2018. Age range: 23–49 years. Denmark;[19]Gallo et al., 2017. Age range: 25–64 years. Italy;[20]Leinonen et al., 2017. Age range: 25–69 years. Norway;[21]Møen et al., 2017. Age range: 25–69 years. Norway;[22]Marlow et al., 2015. Age range: 30–60 years. England.

**Supplementary Figure S2.** Summary of studies on the determinants of condom use (January 2015–December 2025)

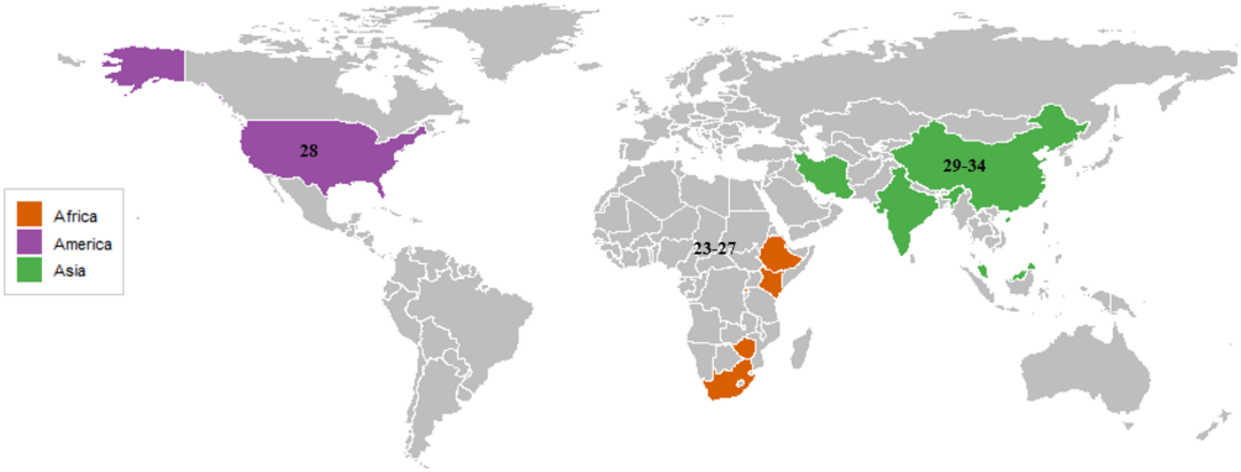


*Note:* This figure presents the countries where at least one study on the topic has been conducted between January 2015 and December 2025, using colour codes by continent. The numbers refer to the corresponding bibliographic references commented below, including information about the participants' age range and the country where the study was conducted.

Africa:[23]Rameto et al., 2023. Age range: 15–59 years. Ethiopia;[24]Sithole et al., 2023. Age range: 15–49 years. Rwanda.[25]Beksinska et al, 2022. Age range: 18–45 years. Kenya;[26]Fearon, 2019 et al,. Age range: 18–65 years. Zimbabwe;[27]Slabbert et al,. 2017. Age range: $<25$–$>35$ years. South Africa.

America:[28]Weir et al., 2015. Age range: 18–62 years. USA.

Asia:[29]Cheah et al., 2025. Age range: $\geq 18$ years. Malaysia.[30]Peng et al., 2025. Age range: $\geq 50$ years. China;[31]Zhong et al., 2023. Age range : 40–85 years. China.[32]Challa et al., 2022. Age range: 18–45 years. India;[33]Zaki et al., 2021. Age range: 15–60 years. Lebanon;[34]Khezri et al., 2020. Age range:$\geq 18$ years. Iran.

### BIBLIOGRAPHY

#### References


[1] AROPE. Glossary: At risk of poverty or social exclusion (AROPE), 2019.

[2] Abiyu Ayalew Assefa, Tihun Feleke, Sintayehu Assefa G/Tsadik, Fekadu Degela, Andualem Zenebe, and Geleta Abera. Utilization and associated factors of cervical cancer screening service among eligible women attending maternal health services at Adare General Hospital, Hawassa city, Southern Ethiopia. *Scientific Reports*, 14(1):2774, feb 2024.

[3] Hinsermu Bayu, Yibrah Berhe, Amlaku Mulat, and Amare Alemu. Cervical cancer screening service uptake and associated factors among age eligible women in Mekelle zone, Northern Ethiopia, 2015: A community based study using health belief model. *PLoS ONE*, 11(3):1–13, 2016.

[4] Jissy Cyriac, Gregory D. Jenkins, Brittany A. Strelow, Danielle J. O' Laughlin, Joy N. Stevens, Kathy L. MacLaughlin, and Jane W. Njeru. A cross-sectional analysis of factors associated with cervical cancer screening in a large midwest primary care setting. *BMC Women's Health*, 25(1):204, apr 2025.

[5] Jessica Law, Geneviève Jessiman-Perreault, Amanda Alberga Machado, Linan Xu, Bonnie Chiang, Huiming Yang, Lisa Allen Scott, Rizwan Shahid, Curtis Mabilangan, Alvin Li, Kamala Adhikari, and Gary Teare. Individual-level characteristics and geospatial factors associated with cervical cancer screening participation in Alberta, Canada: a population-based cross-sectional study. *BMC Public Health*, 25, dec 2025.

[6] Aaron Shaykevich and Martha Wojtowycz. Factors Associated with Cervical Cancer Screening for Women in New York State: Analyzing the 2022 NYS BRFSS. *Journal of Women's Health*, jun 2025.

[7] Ho-Jui Tung, Gila Schwarzschild, Nenrot Gopep, and Ming-Chin Yeh. Cervical Cancer Screening After Menopause. *Healthcare*, 13(10):1157, may 2025.

[8] Marie Rachelle Narcisse, Pearl A. McElfish, Emily Hallgren, Natalie Pierre-Joseph, and Holly C. Felix. Non-use and inadequate use of cervical cancer screening among a representative sample of women in the United States. *Frontiers in Public Health*, 12, 2024.

[9] Victor A. Eng, Sean P. David, Shufeng Li, Mina S. Ally, Marcia Stefanick, and Jean Y. Tang. The association between cigarette smoking, cancer screening, and cancer stage: a prospective study of the women's health initiative observational cohort. *BMJ Open*, 10(8):e037945, aug 2020.

[10] Nicole L. Johnson, Katharine J. Head, Susanna Foxworthy Scott, and Gregory D. Zimet. Persistent Disparities in Cervical Cancer Screening Uptake: Knowledge and Sociodemographic Determinants of Papanicolaou and Human Papillomavirus Testing Among Women in the United States. *Public Health Reports*, 135(4):483–491, 2020.

[11] Cecilia Chinemerem Nwabichie, Rosliza Abdul Manaf, and Suriani Binti Ismail. Factors affecting uptake of cervical cancer screening among African Women in Klang Valley, Malaysia. *Asian Pacific Journal of Cancer Prevention*, 19(3):825–831, 2018.

[12] Marco Ciziceno, Alessia Bertolazzi, and Valeria Quaglia. Examining cervical cancer screening adherence: how does healthism influence participation? *BMC Public Health*, 25(1):3420, oct 2025.

[13] Catarina Gomes Madeira, Claudia Marotta, Anna Georgina Ditter, and Mario Carlo Battista Raviglione. Participation in cervical cancer screening among migrants and non- migrants in primary healthcare in Lisbon : a register- based study. *BMJ Global Health*, 10(12):e019061, 2025.

[14] Silvia Portero de la Cruz and Jesús Cebrino. Trends and Determinants in Uptake of Cervical Cancer Screening in Spain: An Analysis of National Surveys from 2017 and 2020. *Cancers*, 14(10), 2022.

[15] Patrícia Marques, Mariana Geraldes, Ana Gama, Bruno Heleno, and Sónia Dias. Non-attendance in cervical cancer screening among migrant women in Portugal: A cross-sectional study. *Women's Health*, 18:1–9, jan 2022.

[16] María Barrera-Castillo, Rosario Fernández-Peña, María del Olivo del Valle-Gómez, Ana Fernández-Feito, and Alberto Lana. Integración social y cribado del cáncer ginecológico de las mujeres inmigrantes en España. *Gaceta Sanitaria*, 34(5):468–473, 2020.

[17] Elise Harder, Rasmus Hertzum-Larsen, Kirsten Frederiksen, Susanne K. Kjær, and Louise T. Thomsen. Non-participation in cervical cancer screening according to health, lifestyle and sexual behavior: A population-based study of nearly 15,000 Danish women aged 23–45 years. *Preventive Medicine*, 137(October 2019):106119, 2020.

[18] Elise Harder, Kirsten E. Juul, Signe M. Jensen, Louise T. Thomsen, Kirsten Frederiksen, and Susanne K. Kjaer. Factors associated with non-participation in cervical cancer screening – A nationwide study of nearly half a million women in Denmark. *Preventive Medicine*, 111(February):94–100, 2018.

[19] Federica Gallo, Adele Caprioglio, Roberta Castagno, Guglielmo Ronco, Nereo Segnan, and Livia Giordano. Inequalities in cervical cancer screening utilisation and results: A comparison between

Italian natives and immigrants from disadvantaged countries. *Health Policy*, 121(10):1072–1078, 2017.

[20] Maarit K. Leinonen, Suzanne Campbell, Ole Klungsøyr, Stefan Lönnberg, Bo T. Hansen, and Mari Nygård. Personal and provider level factors influence participation to cervical cancer screening: A retrospective register-based study of 1.3 million women in Norway. *Preventive Medicine*, 94:31–39, 2017.

[21] Kathy A. Møen, Bernadette Kumar, Samera Qureshi, and Esperanza Diaz. Differences in cervical cancer screening between immigrants and nonimmigrants in Norway: A primary healthcare register-based study. *European Journal of Cancer Prevention*, 26(6):521–527, 2017.

[22] L A V Marlow, J. Wardle, and J. Waller. Understanding cervical screening non-attendance among ethnic minority women in England. *British Journal of Cancer*, 113(5):833–839, sep 2015.

[23] Muhammed Ahmed Rameto, Saro Abdella, Jemal Ayalew, Masresha Tessema, Jaleta Bulti, Fayiso Bati, and Sileshi Lulseged. Prevalence and factors associated with inconsistent condom use among female sex workers in Ethiopia: findings from the national biobehavioral survey, 2020. *BMC Public Health*, 23(1):2407, dec 2023.

[24] Mkhombiseni Zamani Sithole, Jesca Mercy Batidzirai, Ashenafi Argaw Yirga, and Alfred Musekiwa. Prevalence and factors associated with condom use among women aged 15-49 years in Rwanda using a survey logistic regression model: evidence from the 2019/20 Rwanda Demographic and Health Survey. *Pan African Medical Journal*, 46(121), 2023.

[25] Alicja Beksinska, Emily Nyariki, Rhoda Kabuti, Mary Kungu, Hellen Babu, Pooja Shah, Chrispo Nyabuto, Monica Okumu, Anne Mahero, Pauline Ngurukiri, Zaina Jama, Erastus Irungu, Wendy Adhiambo, Peter Muthoga, Rupert Kaul, Janet Seeley, Helen A. Weiss, Joshua Kimani, and Tara S. Beattie. Harmful Alcohol and Drug Use Is Associated with Syndemic Risk Factors among Female Sex Workers in Nairobi, Kenya. *International Journal of Environmental Research and Public Health*, 19(12):7294, jun 2022.

[26] Elizabeth Fearon, Andrew Phillips, Sibongile Mtetwa, Sungai T. Chabata, Phillis Mushati, Valentina Cambiano, Joanna Busza, Sue Napierala, Bernadette Hensen, Stefan Baral, Sharon S. Weir, Brian Rice, Frances M. Cowan, and James R. Hargreaves. How Can Programs Better Support Female Sex Workers to Avoid HIV Infection in Zimbabwe? A Prevention Cascade Analysis. *JAIDS Journal of Acquired Immune Deficiency Syndromes*, 81(1):24–35, may 2019.

[27] Mariette Slabbert, Francois Venter, Cynthia Gay, Corine Roelofsen, Samanta Lalla-Edward, and Helen Rees. Sexual and reproductive health outcomes among female sex workers in Johannesburg and Pretoria, South Africa: Recommendations for public health programmes. *BMC Public Health*, 17(S3):442, jul 2017.

[28] Brian W. Weir and Carl A. Latkin. Alcohol, Intercourse, and Condom Use Among Women Recently Involved in the Criminal Justice System: Findings from Integrated Global-Frequency and Event-Level Methods. *AIDS and Behavior*, 19(6):1048–1060, 2015.

[29] Yong Kang Cheah, Anita Suleiman, and Mazliza Ramly. Socioeconomic and demographic factors associated with condom use among female sex workers in Malaysia. *Malaysian Journal of Public Health Medicine*, 25(1):181–189, 2025.

[30] Xin Peng, Bingyi Wang, Yong Lu, Xinyi Li, Yuwei Li, Lin Ouyang, Guohui Wu, Yong Cai, Maohe Yu, Jiewei Liu, Yoshiko Sakuma, Hayley Conyers, Xiaojun Meng, Weiming Tang, Joseph D. Tucker, Dan Wu, and Huachun Zou. PrEP-eligible behaviours and condom use among sexually active older adults in China: Findings from the sexual well-being (SWELL) study. *Public Health*, 241(September 2024):164–170, apr 2025.

[31] Xueyuan Zhong, Shuying Chen, Hong Xiao, Xueling Xiao, Simin Yu, Yan Shen, Chen Chen, and Honghong Wang. Perceived HIV risk and factors associated with condom use among women aged 40 and older: A cross-sectional survey. *International Journal of Nursing Sciences*, 10(4):533–539, oct 2023.

[32] Apoorva Challa, Garima Kachhawa, Seema Sood, Ashish D. Upadhyay, Sada N. Dwivedi, and Somesh Gupta. Correlates of bacterial vaginosis among women from North India. *International Journal of STD & AIDS*, 33(7):666–671, jun 2022.

[33] Sara Abu Zaki, Jihane Naous, Antoine Ghanem, Diana Abou Abbas, Roland Tomb, Jade Ghosn, and Ayman Assi. Prevalence of STIs, sexual practices and substance use among 2083 sexually active unmarried women in Lebanon. *Scientific Reports*, 11(1):1–11, 2021.

[34] Mehrdad Khezri, Mostafa Shokoohi, Ali Mirzazadeh, Mohammad Karamouzian, Hamid Sharifi, AliAkbar Haghdoost, and Stefan D. Baral. Early sex work initiation and its association with condomless sex and sexually transmitted infections among female sex workers in Iran. *International Journal of STD & AIDS*, 31(7):671–679, jun 2020.
